# Use of statins and other lipid-modifying agents across pregnancy – a nationwide drug utilization study of 822 071 pregnancies in Norway in 2005-2018

**DOI:** 10.1101/2022.06.28.22276237

**Authors:** Jacob J. Christensen, Martin P. Bogsrud, Kirsten B. Holven, Kjetil Retterstøl, Marit B. Veierød, Hedvig Nordeng

**Author notes:** **Correspondence:** Jacob J. Christensen, PhD. **Disclaimers:** None.

## Abstract

**Background:** Statins are becoming more widely used among women of reproductive age; however, nationwide data on statin use across pregnancy is scarce. We therefore aimed to describe the drug utilization patterns for statins and other lipid-modifying agents (LMAs) before, during, and after pregnancy, for all pregnancies in Norway from 2005 to 2018.

**Methods:** We linked individual-level data from four nationwide electronic health care registries in Norway and characterized the prescription fills of statins and other LMAs across pregnancy. We also examined trends in pregnancy-related LMA use, and characterized women using statins and other LMAs on parameters of health status and co-morbidity.

**Results:** In total 822 071 pregnancies for 503 723 women were included. The number of statin prescription fills decreased rapidly during first trimester and returned to pre-pregnancy levels about one year postpartum. Pregnancy-related statin use increased from 2005 (approx. 0.11 % of all pregnancies) to 2018 (approx. 0.29 % of all pregnancies); however, in total, few statin prescriptions were filled within any trimester of pregnancy (n = 331, 0.04 % of all pregnancies). Statin use was more common in women with higher age, higher weight, smoking, and comorbidities such as hypertension and diabetes mellitus; also, statin users often had co-medication pertinent to these conditions.

**Conclusions:** Although statins and other LMAs were increasingly being used around the time of pregnancy among women in Norway, drug use was mostly discontinued during first trimester. Our results suggest that pregnancy-related statin use should be monitored, and that drug safety analyses for maternal and offspring health outcomes are needed.

## Introduction

Statins (HMG coenzyme A [HMG-CoA] reductase inhibitors) reduce plasma LDL cholesterol (LDL-C) and thereby the risk of atherosclerotic cardiovascular disease (ASCVD) both effectively and safely [1, 2]. Statins therefore constitute the main line of therapy for patients with elevated ASCVD risk [3–5]. In recent years, clinical guidelines have further lowered the threshold for initiating treatment with statins [3–5]; thus, statins are now more commonly used among the general population [6]. Drug Wholesales Statistics in Norway show that statin use has increased over time also among women of reproductive age [7]. In 2004, 1 in 182 women aged 20-44 years in Norway prescribed at least one lipid-modifying agent (LMA, ATC [Anatomical Therapeutic Chemical Classification] group C10), the majority of which was statin monotherapy (ATC group C10AA). In 2020, the rate of LMA use had increased to 1 in 126 women aged 20-44 years.

We may expect a further increase in the number of women of reproductive age who are eligible for statin therapy in the coming years. The reason is at least two-fold. First, as hypercholesterolemia is being detected more frequently and earlier in life, and the threshold for initiating treatment is lowered, so is the age that statin therapy is started. This is a natural consequence of the realization that *duration of treatment* (number of treatment years) is key to effective risk reduction [8]. Second, the average maternal age at first pregnancy is still increasing, leading to a higher proportion of women of reproductive age with elevated ASCVD risk [9]. Importantly, considering that a proportion of all pregnancies are unplanned, the expected increase in statin use will probably affect the population-wide exposure to statins also in early pregnancy. However, statins are *contraindicated* during pregnancy and lactation, due to proposed teratogenic effects and lack of safety data. Women planning pregnancy are therefore recommended to stop statin therapy [4].

Although important both from a cardioprotective and safety point of view, nationwide data on statin use across pregnancy is scarce. Therefore, in the present study, we aimed to describe the complex drug utilization patterns for statins and other lipid-modifying agents (LMAs) before, during, and after pregnancy, for all pregnancies in Norway in the period 2005 to 2018, using nationwide linked electronic health care registries. A secondary aim was to determine health-related characteristics of women using statins and other LMAs during pregnancy.

## Material and methods

### Study design, data sources and study population

The present study was a nationwide registry drug utilization study where we described the use of statins and other LMAs across pregnancy for all pregnancies in Norway in the period 2005 to 2018.

Using the unique, personal identity number of Norwegian citizens, we linked individual-level data from four national health registries in Norway up until 2018: The Medical Birth Registry of Norway (MBRN), the Norwegian Prescription Registry (NorPD), the Norwegian Patient Registry (NPR), and the Norway Control and Payment of Health Reimbursement Database (KUHR).

The MBRN has registered pregnancy-related information since 1967, including pregnancy duration, complications, growth of the fetus, and health status of both the mother and the child, for all pregnancies in Norway ending after gestational week 12 [10]. NorPD has recorded all prescribed medications dispensed at Norwegian pharmacies to non-institutionalized individuals since 2004, including the type of drug, the date of prescription filling, and the number of defined daily doses (DDDs) dispensed [11]. Medications in NorPD are classified according to the World Health Organization’s Anatomical Therapeutic Chemical Classification (ATC) System (see https://www.whocc.no/ for details).

NPR includes information on activity in both secondary and tertiary health care settings in Norway. The NPR uses the International Classification of Diseases, version 10 (ICD-10). KUHR stores information on activity in *primary* health care settings in Norway. KUHR records the reason for the utilization of health care services, which includes visits to general practitioners (GPs) and emergency services. KUHR uses both ICD-10 diagnostic codes and the International Classification of Primary Care-2 (ICPC-2) codes.

For the present study, we included all women with a pregnancy recorded in the MBRN in 2005-2018. We excluded pregnancies with missing gestational length, as timing of drug exposure could not be calculated for these pregnancies (**Figure 1**). Pregnancies where women used statins and other LMAs were identified through dispensed prescriptions in NorPD.

**Figure 1.**
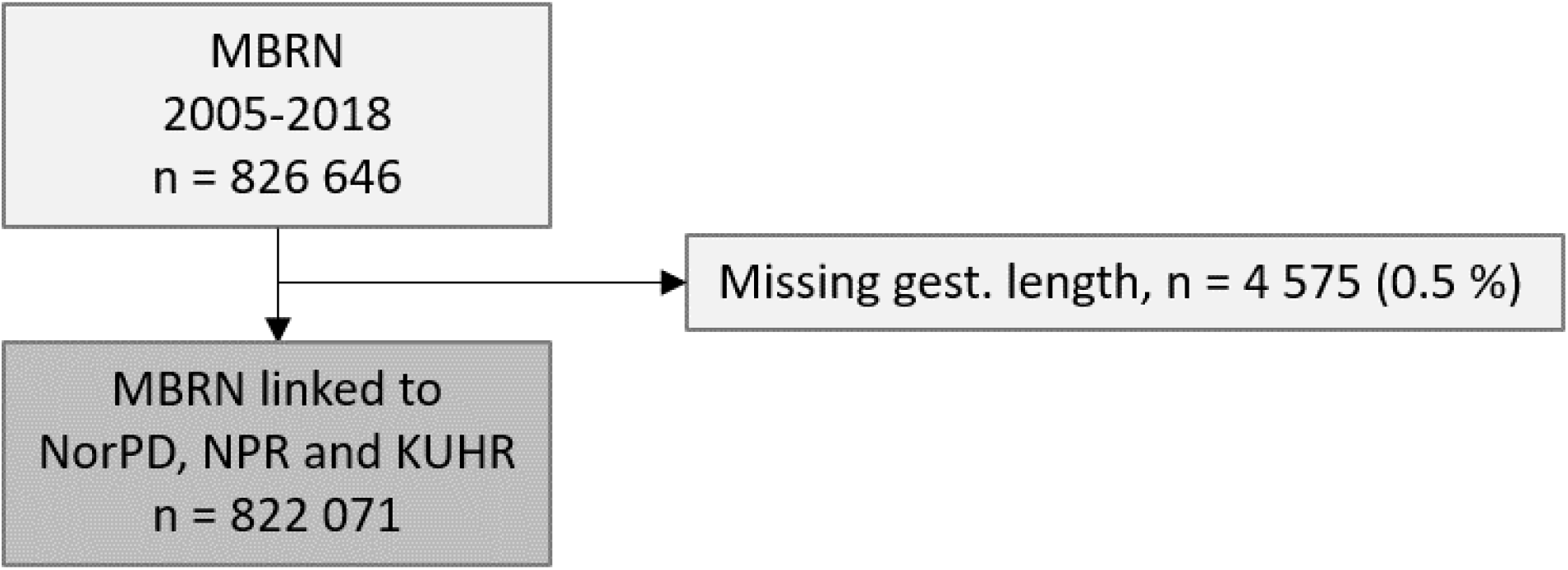
Study flow chart. Abbreviations: gest, gestational; KUHR, the Norway Control and Payment of Health Reimbursement Database; MBRN, the Medical Birth Registry of Norway; NorPD, the Norwegian Prescription Registry; NPR, the Norwegian Patient Registry.

To characterize women using LMAs during pregnancy, we classified pregnancies into different subgroups based on different criteria. First, limited to one year before conception to one year after delivery, we classified pregnancies into the following groups:

- Classification no. 1 (statin groups): Pregnancies for women using any statin or specific statins subtypes.
- Classification no. 2 (non-statin LMA groups): Pregnancies for women using any non-statin LMA, or specific non-statin LMA subtypes.

Similarly, limited to any time before conception to one year after delivery, we classified pregnancies into the following groups:

- Classification no. 3 (“discontinuation” groups): 1) pregnancies where women discontinued their statin prescription fills at least three months before conception, 2) pregnancies where women discontinued their statin prescription fills within three months before conception, 3) pregnancies where women filled their statin prescription during pregnancy, and 4) other.
- Classification no. 4 (groups based on history of statin use before pregnancy): 1) pregnancies for long-term, “experienced” statin users (10-148 prescription fills), 2) pregnancies for “intermediate-term” users (4-9 prescription fills), and 3) pregnancies for “short-term users and beginners” (1-3 prescription fills). History of statin use was calculated as the chronological, rolling count of prescription fills per woman and pregnancy. We determined the ranges by binning the number of prescriptions into three groups of approximately equal number of observations.
- Classification no. 5 (dyslipidemia diagnosis groups): 1) pregnancies for women with a dyslipidemia diagnosis (ICPC-2 code T93) not using LMAs, and 2) pregnancies for women with a dyslipidemia diagnosis using LMAs.
- Classification no. 6 (co-morbidity groups based on number of indications for LMA therapy): pregnancies for women using LMA with 1) 0 indications for LMA use, 2) 1 indication for LMA use, 3) 2-3 indications for LMA use, 4) 4-8 indications for LMA use, or 5) ≥9 indications for LMA use. Number of LMA indications was defined as the per-pregnancy sum of unique drugs used in diabetes (ATC code A10), antithrombotic agents (B01), and cardiovascular system (C other than C10), and the unique diagnosis codes defined in **Table S1**.

This project was approved by the Regional Committee for Research Ethics in South-Eastern Norway (approval number 2018/140/REK South-East) and by the Data Protection Officer at the University of Oslo (approval number 58033).

### Exposure to statins and other LMAs

Exposure to statins and other LMAs was defined based on the date of prescription dispensing and the dispensed amount expressed in number of DDDs. We assumed that all the dispensed statins were consumed as 1 DDD per day. The main exposures of interest were prescription fills of statins, as well as prescription fills of other, non-statin LMAs. The ATC codes for statins include C10AA01 (simvastatin), C10AA05 (atorvastatin), and C10AA07 (rosuvastatin). The ATC codes for other LMA included C10AC01 (colestyramine), C10AX06 (omega-3 triglycerides incl. other esters and acids), and C10AX09 (ezetimibe).

We defined exposure as the presence of at least one prescription fill within each time frame. Time frames of interest were mainly between one year before conception to one year after delivery, including the entire pregnancy and pregnancy trimesters (trimester 1: 1-93 days after time of conception; trimester 2: 94-186 days after conception; trimester 3: > 186 days after conception).

### Co-medication

We examined co-medication among women using statins and other LMAs. Among co-medication where LMAs would likely be indicated, we included drugs used in diabetes (ATC code A10), antithrombotic agents (B01), and cardiovascular system (C other than C10). We also had access to a range of other co-medications which we examined using the first ATC level, including alimentary tract and metabolism (A), blood and blood-forming organs (B), genito-urinary system and sex hormones (G), systemic hormonal preparations excl. sex hormones and insulins (H), antiinfectives for systemic use (J), musculo-skeletal system (M), and nervous system (N). Where relevant, we also considered co-medications at the second ATC level. We omitted some ATC codes that had either few observations or were considered less relevant, such as dermatologicals (D), antineoplastic and immunomodulating agents (L), antiparasitic products, insecticides and repellents (P), respiratory system (R), sensory organs (S), and various (V).

### Diagnoses

We examined diagnoses among women using statins and other LMAs using ICD-10 codes in NPR and ICD-10 and ICPC-2 codes in KUHR. Among diagnostic codes where LMAs would likely be indicated, we included 28 second level ICD-10 codes from endocrine, nutrition and metabolic (ICD-10 main group E) and circulatory (I) diagnostic groups; we also included 22 second level ICPC-2 codes from the cardiovascular (K) and endocrine, metabolic and nutrition (T) (Table S1). Where relevant, we highlighted aggregated subgroups, such as T9, which cover diabetes type 2 (ICPC-2 code T90) and dyslipidemias (T93). Notably, we did not have access to some relevant diagnosis such as metabolic disorders (E70-E90) which include dyslipidemias (E78) and the subgroup familial hypercholesterolemia (E78.0).

We also had access to a range of other diagnoses, including the following ICD-10 codes: psychiatric and behavior (F), nervous system (G), pregnancy, birth and maternity (O), and miscellaneous (Z); for the ICPC-2 codes, we included musculoskeletal (L), nervous system (N), psychiatric (P), airways (R), urinary tract (U), and pregnancy, birth and family (W). We omitted some diagnosis codes that had either few observations or were considered less relevant, such as skin (L) and respiratory (J) for ICD-10, and social (Z) for ICPC-2.

### Maternal characteristics

We derived the following information from the MBRN: maternal age at delivery, marital status, parity, pre-pregnancy body weight, employment, pre-pregnant folate use, smoking, comorbidities (asthma, chronic hypertension, chronic renal disease, pre-existing diabetes, epilepsy, and rheumatoid arthritis), plurality, previous pregnancy loss, and an obstetric comorbidity index (adapted from [12]). Variables were categorized as presented in the tables and figures.

### Recurrent pregnancies

Recurrent pregnancy was defined as any record of a pregnancy ending after gestational week 12, including both live and non-live births (that is, elective terminations, late miscarriages after gestational week 12, and stillbirths).

### Data analyses

We estimated the number of prescription fills for statins and other LMAs from one year before conception, during pregnancy, and up to one year after delivery. We also estimated the time trends of use from 2005 to 2018 as a proportion of total number of pregnancies per year. We used several visualization approaches to display the results, for example using the number of prescription fills per 2-week period as a function of time (using estimated time of conception as time zero). See further details in each figure legend.

Using descriptive statistics, we evaluated the main characteristics, comorbidities, and concomitant medications of women with and without LMA prescription fills. For example, in tables, we present frequencies (%) and medians (interquartile ranges, IQR).

All data analyses were performed in R version 3.6.2 [13] using RStudio (Boston, MA, USA, www.rstudio.com) and the tidyverse framework [14].

## Results

We included 822 071 pregnancies for 503 723 unique women (Figure 1). Half of the women contributed with one pregnancy (n = 250 664, 49.8 %), and a sizable proportion contributed with two (n = 196 579, 39.0 %) or three pregnancies (n = 49 119, 9.8 %); few women contributed with four or more pregnancies (n = 7 361, 1.5 %). The characteristics of the pregnancies is shown in **Table 1**. Compared to non-statin users, statin users had higher maternal age (47 vs. 31 % above 32 years for any statin use and no statin use, respectively), higher pre-pregnancy maternal weight (70 vs. 65 kg), higher prevalence of smoking (25 vs. 19 %), and comorbidities, such as any chronic illness (24 vs. 6.7 %), chronic hypertension (6.5 vs. 0.5 %), or diabetes (18 vs. 3.5 %) (Table 1). This pattern was consistent from 2005 to 2018 (**Figure S1**).

**Table 1.**
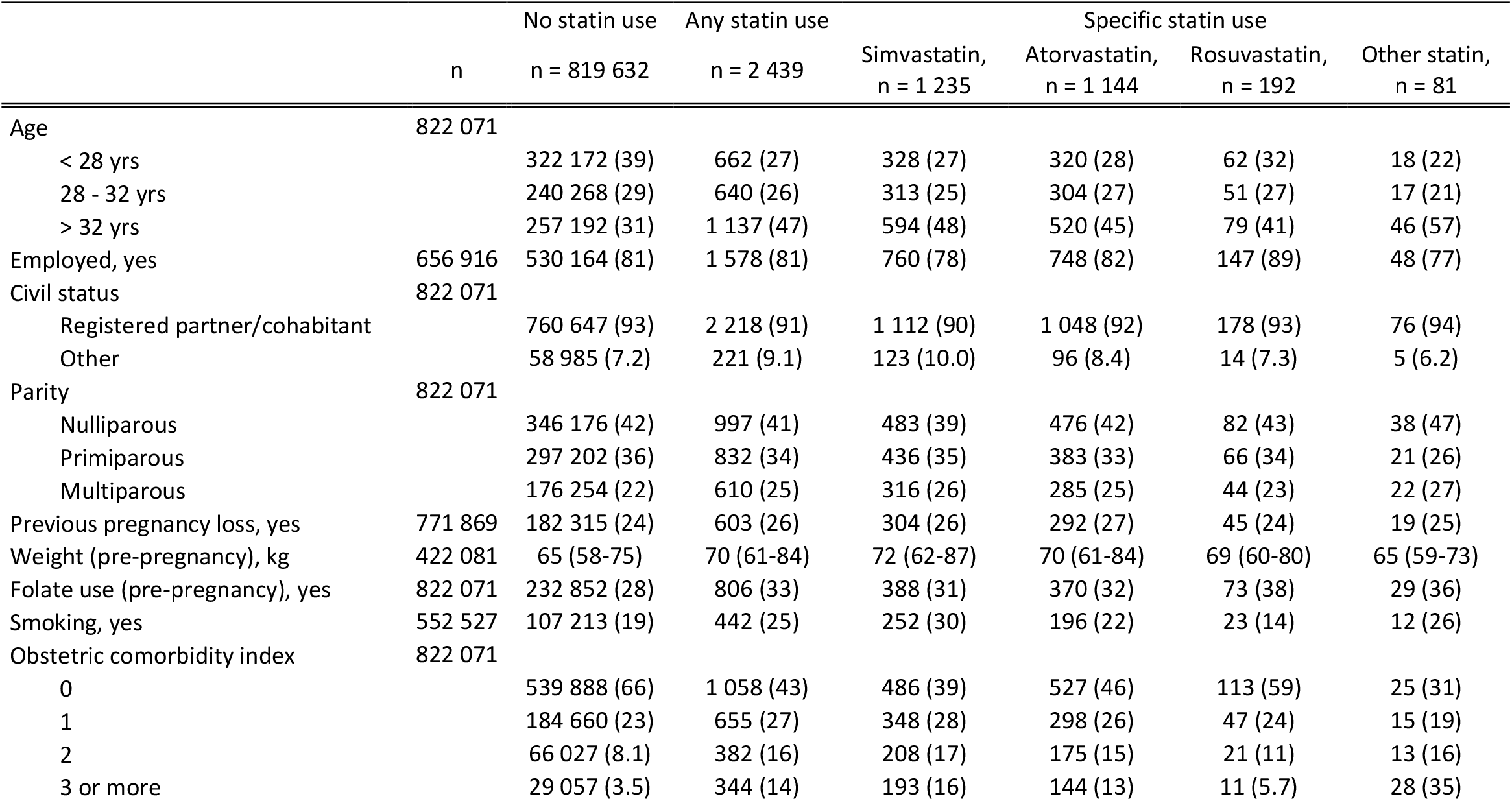
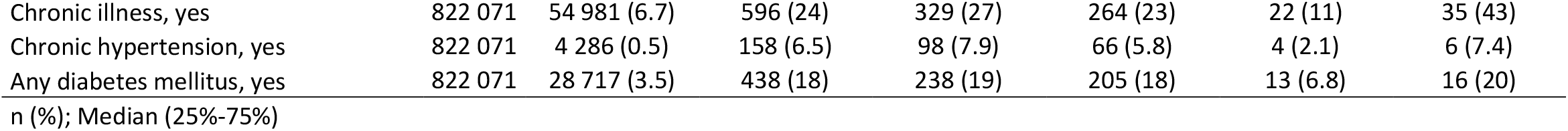
Characteristics of the 822 071 pregnancies in 2005-2018, stratified by statin use between one year before conception and one year after delivery (covering 2 439 pregnancies and 2 010 unique women).

### LMA use across pregnancy

In the period 2005 to 2018, between one year before conception and one year after delivery, 8 255 prescriptions were filled for an LMA, covering 2 772 pregnancies and 2 304 unique women; of these, 6 703 prescription fills (81.2 %) were for a statin, covering 2 439 pregnancies and 2 010 unique women (**Figure 2**). The main statin types used were simvastatin and atorvastatin. Although the number of prescriptions decreased during the first trimester of pregnancy, statin prescriptions were filled in first trimester for a total of 298 unique pregnancies (0.036 % of all pregnancies), and in any trimester for a total of 331 unique pregnancies (0.04 % of all pregnancies). The reduction in prescription fills started already three months before pregnancy, from almost 150 to about 100 prescription fills per two-week period. The number of prescription fills remained low throughout second and third trimester (less than 10 prescription fills per two-week period), increased steadily following pregnancy, and returned to pre-pregnancy levels after almost one year postpartum. The pattern was similar for all statin types and doses (**Figure S2**).

**Figure 2.**
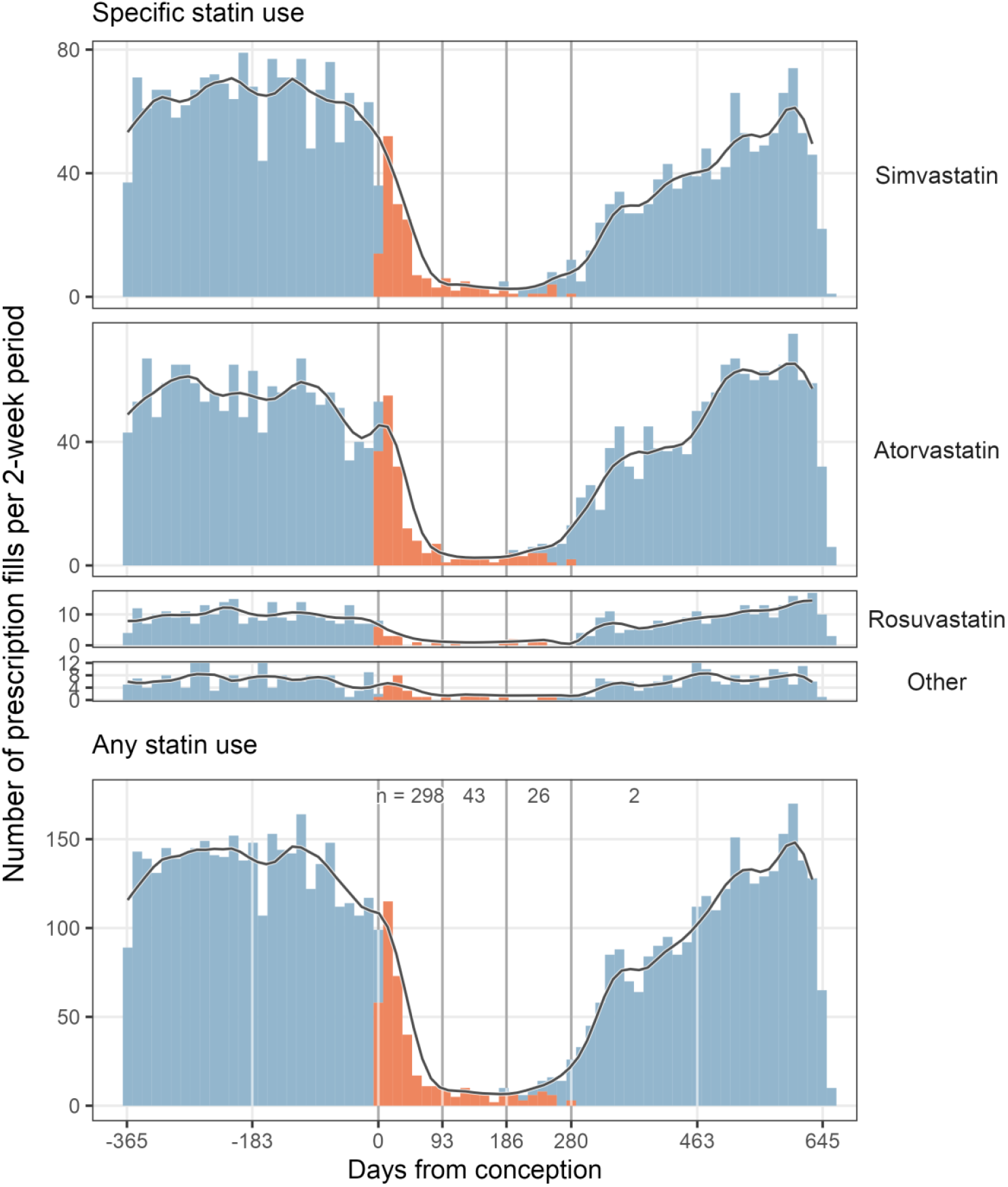
Pattern of prescription fills for statins, one year before, during, and one year after pregnancy. The figure shows the number of prescriptions fills for statins (6 703 prescription fills, 2 439 pregnancies, 2 010 unique women) in 2-week bins, as well as a smoothened 2-month average (one month prior and the subsequent two weeks; local regressions were fit on 10 % of the data points along the line). Blue and red color represents prescription fill of a statin *outside* or *within* any trimester of pregnancy, respectively. Numbers between the vertical lines in the upper part of the lowest panel represent the number of unique pregnancies where a statin prescription had been filled inside pregnancy, per trimester. The vertical lines represent key timepoints in relation to time of conception, including conception and trimester transitions.

Non-statin LMAs were less commonly used across pregnancy compared with statins. In the period 2005 to 2018, between one year before conception and one year after delivery, 1 659 prescriptions were filled for a non-statin LMA, covering 599 pregnancies and 519 unique women (**Figure S3**). While ezetimibe use followed a pattern similar to that of statins, the use of cholestyramine and omega-3 was more stable across pregnancy. Non-statin LMA prescriptions were filled in first trimester for a total of 89 unique pregnancies (0.011 % of all pregnancies), and in any trimester for a total of 121 unique pregnancies (0.015 % of all pregnancies).

### Temporal trends in pregnancy-related LMA use

Pregnancy-related LMA use increased from 2005 to 2018, both for statins and other LMAs (**Figure 3**). Within 1 year before pregnancy, any LMA use increased from 0.14 % of all pregnancies in 2006 to 0.29 % of all pregnancies in 2018; similarly, within 1 year after pregnancy, any LMA use increased from 0.11 % of all pregnancies in 2005 to 0.23 % of all pregnancies in 2017. In contrast, any LMA use during pregnancy was approximately 0.05 % of all pregnancies throughout the whole period. Simvastatin was the most frequently used drug until approximately 2013, when atorvastatin (and rosuvastatin) became more frequently used. Use of ezetimibe and colestyramine also increased over time.

**Figure 3.**
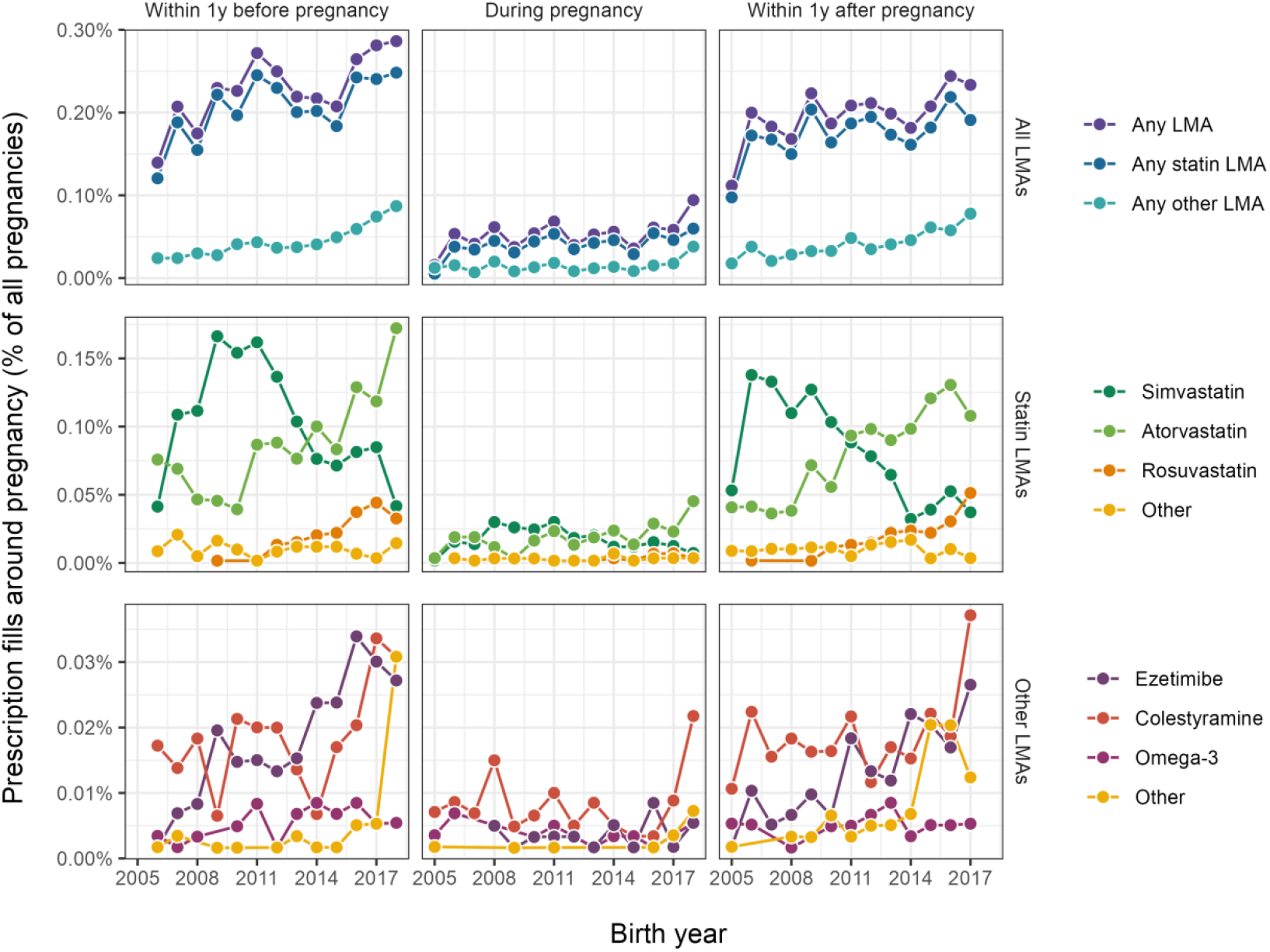
Trends in prescription fills for LMAs, one year before, during, and one year after pregnancy. The figure shows trends in use for groups of LMAs (top row) and individual LMAs (middle and bottom rows), for the periods from 1 year before pregnancy to conception (left column), during pregnancy (middle column), and from delivery until 1 year after delivery (right column). Note that 2005 was excluded for the period within 1 year before pregnancy, as this would largely reflect 2004; similarly, 2018 was excluded for the period within year after pregnancy, as this would largely reflect 2019. Abbreviations: LMA, lipid-modifying agent; y, year.

### Co-medication across pregnancy

Women using LMAs were using cardiovascular co-medication across pregnancy (**Figure 4**). For example, co-medication with alimentary tract and metabolism drugs (ATC group A) increased in pregnancy, partly resulting from diabetes drugs (A10). Similarly, drugs for blood and blood-forming organs (B) increased during pregnancy, mainly as a result of anti-thrombotic agents (B01). Also, certain subgroups of cardiovascular system drugs (C) increased slightly towards the end of pregnancy and in the immediate time following delivery, such as vasoprotectives (C05), beta-blockers (C07), calcium channel blockers (C08), and renin-angiotensin system (C09).

**Figure 4.**
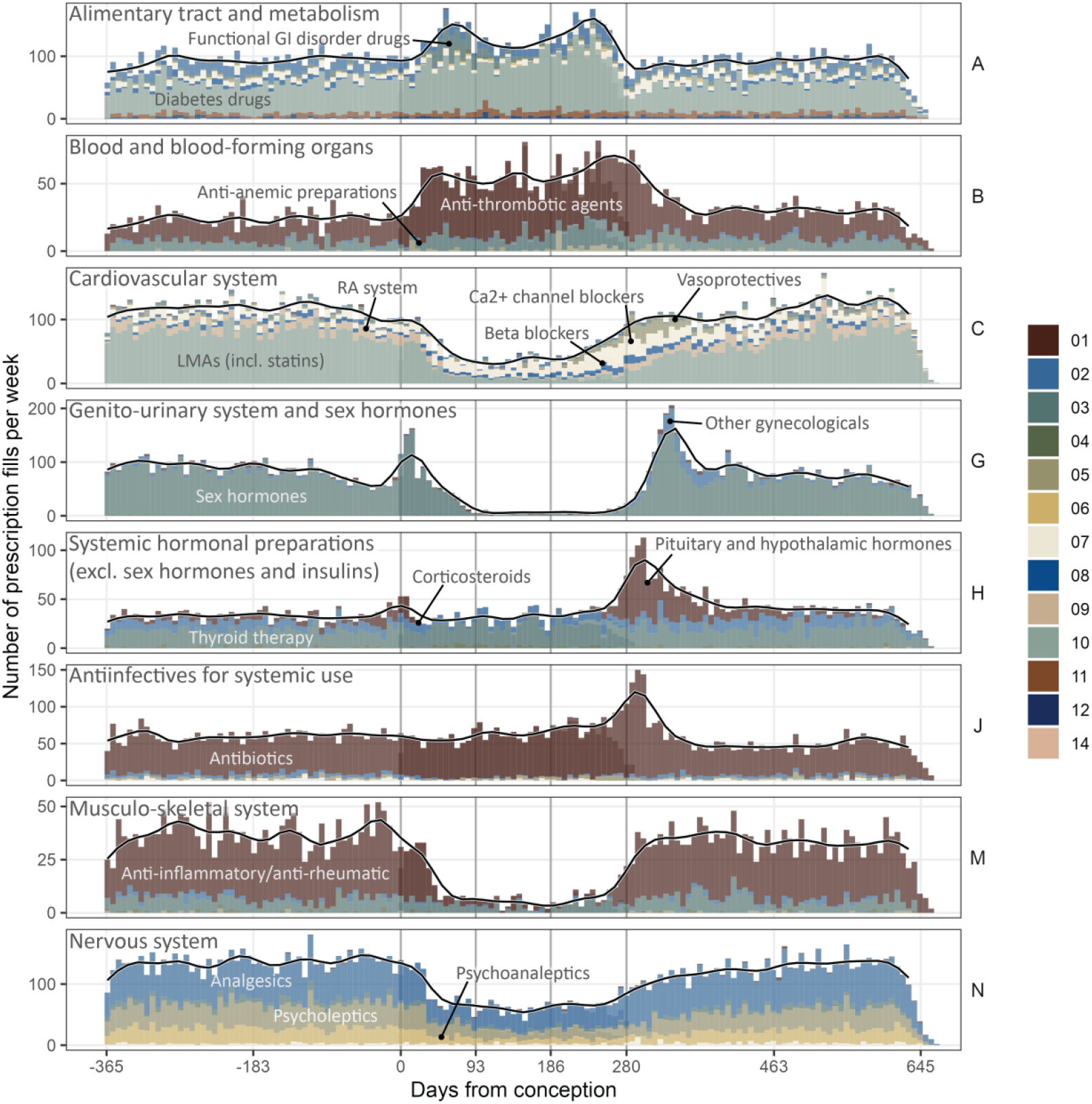
Pattern of co-medication among LMA users one year before, during, and one year after pregnancy. The figure shows the number of prescriptions fills of various medication in 1-week bins for women using statins across pregnancy (those included in Figure 2, covering 2 439 pregnancies and 2 010 unique women), as well as a smoothened 1-month average (two weeks prior and the subsequent week; local regressions were fit on 10 % of the data points along the line). The panels and colors correspond to the first and second level of ATC codes, respectively. Some of the most notable second level ATC codes are annotated by their common names. The vertical lines represent key timepoints in relation to time of conception, including conception and trimester transitions..

Approximately 37 % of women who used LMAs in pregnancies also used a cardiovascular system drug *other* than LMAs, including antihypertensives, and 16 % used diabetes drugs (data not shown).

### Diagnoses across pregnancy

Women using LMAs were being diagnosed with conditions across pregnancy pertinent to LMA use (**Figure 5**). For example, group E1 in ICD-10, covering endocrine, nutrition and metabolic diseases including diabetes spiked during pregnancy; however, T9 in ICPC-2, which include diabetes mellitus type 2 and dyslipidemia, did not. Group I in ICD-10 (circulatory) increased towards the end of pregnancy and following delivery, while group K in ICPC-2 (cardiovascular) increased following delivery. We counted at least one diagnosis of dyslipidemia (ICPC-2 code T93 in KUHR) for 62 % of all pregnancies where a prescription for an LMA had been filled between one year before conception and one year following delivery (**Figure S4**).

**Figure 5.**
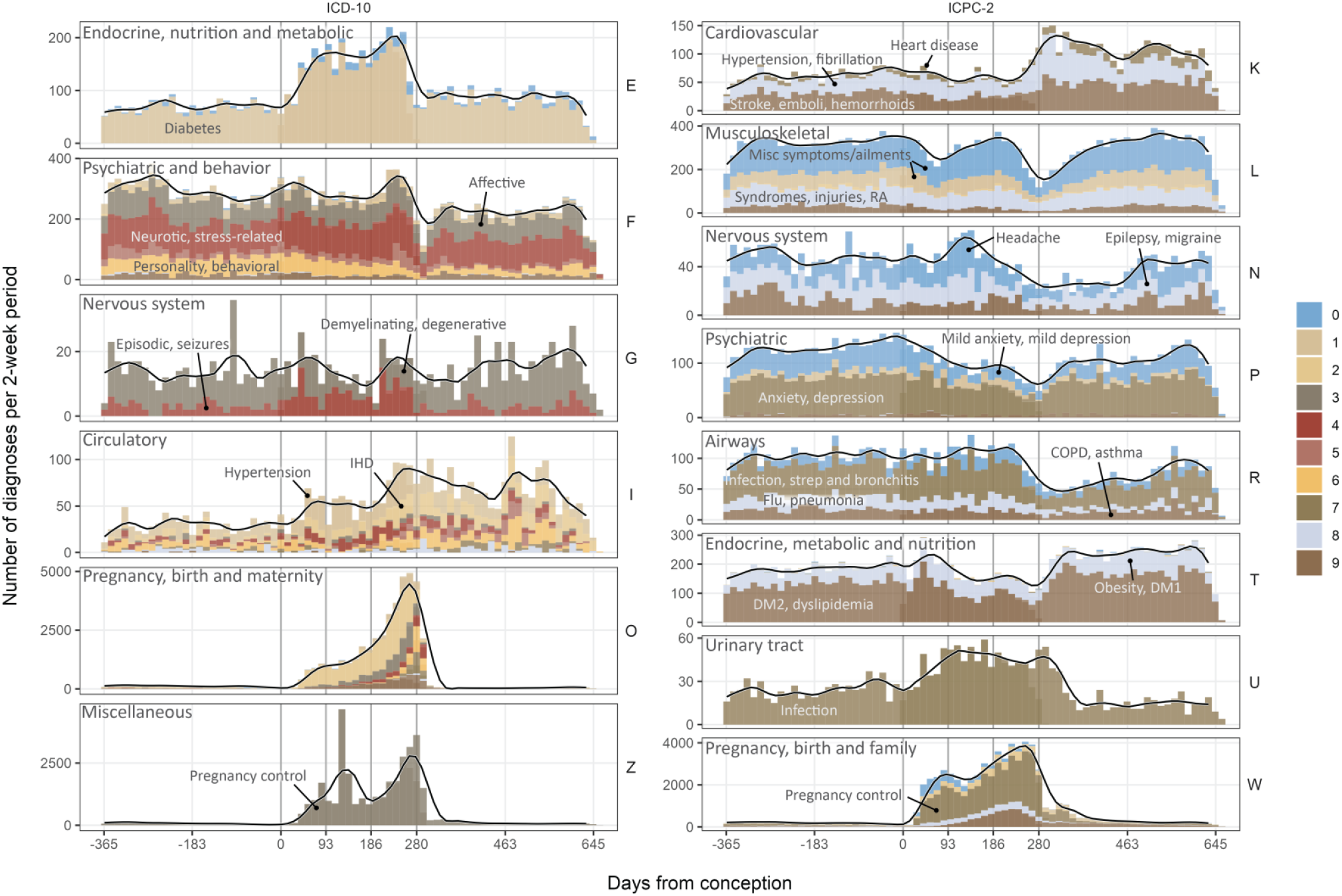
Pattern of the most common diagnoses given among LMA users one year before, during, and one year after pregnancy. The figure shows the number of diagnoses in 2-week bins for women using statins across pregnancy (those included in Figure 2, covering 2 439 pregnancies and 2 010 unique women), as well as a smoothened 2-month average (one month prior and the subsequent two weeks; local regressions were fit on 10 % of the data points along the line). The count for ICD-10 was based on either NPR or KUHR. The data was not restricted to first occurrence of a diagnosis; hence, each woman can be represented by the same diagnosis multiple times. The panels and colors correspond to the first and second level diagnosis codes, respectively. Some of the most notable second level diagnosis codes are annotated by keywords reflecting broad sets of diagnosis terms. The vertical lines represent key timepoints in relation to time of conception, including conception and trimester transitions.

### Characteristics of pregnancies based on additional subgroups

In the Supplementary Results and Discussion, we present and discuss the characteristics of pregnancies for LMA users more in-depth, based on several additional subgroups, including use of non-statin LMAs, and discontinuation (**Table S2-S6, Figure S5-S7**).

## Discussion

In the present work, we explored the use of statins and other LMAs across pregnancy for all pregnancies in Norway in the period 2005 to 2018. We found that statin use displayed a distinct pattern across pregnancy: The number of statin prescription fills decreased during first trimester, which indicates that women discontinue statins once they know they are pregnant, as recommended by guidelines. Furthermore, we found that pregnancy-related LMA use increased from 2005 to 2018, and that women using statins and other LMAs had different characteristics than other women. Our results highlight the importance of monitoring pregnancy-related statin use and should inform drug safety analyses for maternal and offspring health outcomes.

### LMA use displayed a distinct pattern across pregnancy

The number of statin prescription fills decreased during first trimester, indicating that women discontinue statins once they know they are pregnant. In fact, the decrease in statin prescription fills started already three months before pregnancy, likely reflecting pregnancy planning, at least among a subgroup of the population. Also, despite the high number of pregnancies (n = 822 071), in relatively few pregnancies (n = 298, 0.036 %), a prescription for a statin had been filled during first trimester. This is largely in line with data from other populations [15–19]. Although reassuring, the low frequency of statin exposure during pregnancy limits the power of drug safety analyses.

The increase in prescription fills occurred slowly and returned to pre-pregnancy levels about one year postpartum. The slow increase could reflect at least two things: statin contraindication during breastfeeding, and leftover pills from before pregnancy. Notably, there was a trend that beginners started using statins earlier than other women; this group likely comprise women that during pregnancy become aware they are at increased ASCVD risk. These data suggest that, in practice, pregnancy is used by the clinicians to assess ASCVD risk and to initiate treatment, at least for some women. Doing this more systematically for all women could be one way to bridge the gap in ASCVD assessment and treatment for women compared to men, which has recently been highlighted [20, 21]. Also, because pregnancy can be considered a physiological test of the pregnant woman and family, pregnancy-related assessment of ASCVD risk may be a primordial prevention strategy worth considering implemented on a more large-scale level [22, 23].

The pregnancy-induced changes in prescription fills for non-statin LMAs were also remarkable. While prescription fills for ezetimibe followed a pattern similar to that of statins, other non-statin LMAs were less affected by pregnancy. Because there is a lack of safety data for *all* LMAs, guidelines generally recommend that *no* LMAs – statin or non-statin – be dispensed during pregnancy or lactation [4]. However, bile acid sequestrants such as cholestyramine may be considered for women with a severe FH phenotype, since these are not absorbed. Therefore, in the present data, cholestyramine use can potentially be seen as a marker of a severe type of FH in the absence of the actual ICD-10 code for FH (E78.0).

### Pregnancy-related LMA use increased over time

Pregnancy-related LMA use increased from 2005 to 2018. As expected, monotherapy with simvastatin and atorvastatin were the main groups of LMAs, corresponding well with trends in use of statin type in clinical trials [4]. Interestingly, the use of combination therapy increased in the past few years, possibly due to the results from the IMPROVE-IT trial, which confirmed clinical benefit of adding ezetimibe on top of statin therapy for ASCVD prevention [24]. Following new reimbursement policies in June 2005, simvastatin was made the drug of choice in Norway [25]. In accordance with these regulations, many low dose atorvastatin users switched to simvastatin instead, and new users were more often put on simvastatin. Over time, atorvastatin again became more widely used, and is now the drug of choice, likely also because of reimbursement rules and introduction of cheap, generic medications [6].

Rosuvastatin monotherapy also became more popular the last decade, which may be related to improved detection and treatment of subjects with familial hypercholesterolemia (FH). In general, these patterns of statin use correspond well with the pattern of statin use in the whole Norwegian population [6].

### Women using statins and other LMAs across pregnancy had different characteristics than other women

Women using statins and other LMAs across pregnancy had higher pre-pregnancy age and weight, a larger proportion of smokers, and various co-morbidities, such as hypertension, kidney disease and diabetes mellitus; factors that reflect elevated ASCVD risk and thereby indication for treatment with statins [4]. This is in line with previous observations: pregnant women using statins generally have a more metabolic unhealthy phenotype than those not using statins, for example due higher prevalence of diabetes mellitus, smoking, or dyslipidemia, depending on the study population [16, 17]. Scientists have long argued that people with high *lifetime* risk of ASCVD events would likely benefit with statin treatment already early in life, even if the *short-term* risk is negligible [26–28]. For dyslipidemia, evidence from FH is the archetypical example in this regard, although contemporary studies using genetics have facilitated a wider recognition across medical sub-disciplines [27]. However, the lifetime ASCVD risk related to *other* risk factors is also highly relevant, as our data here point to. With time, as the concept of lifetime risk is being adopted by clinicians, more people are likely to be treated at a younger age, including women of reproductive age. It is therefore pivotal to keep pregnancy-related statin use under surveillance.

### Co-medication and diagnoses were common among women using statins and other LMAs, supporting a more prevalent co-morbidity

Interestingly, each pharmacological group and diagnosis group showed very distinct patterns across pregnancy. The use of both anti-thrombotic and anti-diabetic medication increased in first trimester and decreased after delivery. Concomitant use of non-LMA cardiovascular drugs was also common across pregnancy, and for some pharmacological subgroups even increased slightly in third trimester (vasoprotectives, beta-blockers, calcium channel blockers, and renin-angiotensin system). Collectively, these data further suggest that the pregnancies of women using LMAs can be considered high-risk pregnancies. Also, the pronounced co-medication highlights the importance of considering the concurrent use of *other* potentially teratogenic drugs and interaction of these with statins when evaluating statin safety [19].

Interestingly, the use of ICD-10 diagnoses for diabetes mellitus (group E1) was high already before pregnancy among LMA users, and increased midway through first trimester. Prenatal care in Norway and elsewhere focuses on detecting alterations in glucose homeostasis in pregnant women, and for good reason [29]. The HAPO study found that maternal glucose concentration associated with risk of developing several adverse health outcomes, such as pre-eclampsia, preterm delivery and macrosomia [30]. Importantly, in that study, the association was both linear and had no apparent threshold. In addition, gestational diabetes mellitus is an excellent predictor of future development of insulin resistance and diabetes mellitus in both mother and offspring [22, 23]. In contrast, the use of ICPC-2 diagnoses for both diabetes mellitus and dyslipidemia (group T9) went slightly down in pregnancy.

Because pregnancy induces a physiological hyperlipidemia, guidelines for prenatal care do usually not recommend routinely measuring lipids in pregnancy, thereby masking the pattern observed with ICD-10 diagnoses.

### Implications, importance & generalizability

The use of complete data from nationwide registries with mandatory reporting are important strengths of our study. Nationwide data on statin use across pregnancy is scarce, despite the importance both from a cardioprotective and safety point of view. ASCVD is ubiquitous worldwide, and lipid-modification is the primary strategy for ASCVD risk reduction. The present work is therefore likely generalizable to other populations where use of statins and other LMAs is becoming more prevalent also among young people. Also, one implication of our results is that multi-national collaboration projects may be needed to reliably answer pressing research questions on statin safety, for example related to outcomes such as birth defects in the offspring. Our study covers all pregnant women in Norway from 2005 to 2018, as opposed to a prospective cohort study that includes only a sample of a population. Another strength is that the registry data was collected in standardized forms by health personnel, in contrast to observational studies using differing assessment methods, and self-reporting.

### Limitations

This work has some limitations that warrant mention. First, the exposure is ‘prescription fill’, not *actual* drug use; however, we assume that these associate closely [31]. Note that the impact of primary non-adherence is limited by the fact that only information on drugs actually dispensed from pharmacies to patients is recorded in the NorPD, which is more indicative of use than databases including all drugs prescribed by physicians [32]. Second, because this study is a sub-study under a larger project regarding medications in pregnancy, we only had access to certain diagnostic codes; for example, we did not have available the ICD-10 code for FH (E78.0). Third, the data is descriptive, and cannot draw any conclusions about the appropriateness of treatment.

### Conclusions

In conclusion, the use of statins and other LMAs displayed a distinct pattern across pregnancy. Reassuringly, the number of prescription fills decreased during first trimester. Pregnancy-related statin use increased over time, and women using statins and other LMAs had more co-morbidity compared to other women. These results suggest that pregnancy-related statin use should be monitored and emphasize the need for drug safety analyses for maternal and offspring health outcomes. Finally, our data point to a more widespread use of periconceptional counselling to women of childbearing age that may require statins or other LMAs.

## Data Availability

All data produced in the present study are available upon reasonable request to the authors

## Abbreviations

ATC: The World Health Organization–s Anatomical Therapeutic Chemical Classification System
DDDs: defined daily doses
ICD-10: The International Classification of Diseases, version 10
ICPC-2: The International Classification of Primary Care-2
KUHR: The Norway Control and Payment of Health Reimbursement Database
LMA: lipid-modifying agents
MBRN: The Medical Birth Registry of Norway
NorPD: The Norwegian Prescription Registry
NPR: The Norwegian Patient Registry

## Acknowledgements

We would like to thank Sarah Hjorth, Department of Pharmacy at UiO, for pre-processing of the raw registry data.

## Authorship

Authorship overview according to CRediT – Contributor Roles Taxonomy: Conceptualization: JJC, KBH, MPB, HN; Data curation: JJC, HN; Formal analysis: JJC; Funding acquisition: HN, KBH, MPB; Investigation: JJC, HN; Methodology: JJC; Project administration: JJC, KBH, HN; Resources: HN; Software: JJC; Supervision: MPB, KBH, KR, MBV, HN; Validation: JJC; Visualization: JJC; Writing – original draft: JJC; Writing – review & editing: JJC, MPB, KBH, KR, MBV, HN.

## References

1. Collins R, Reith C, Emberson J, Armitage J, Baigent C, Blackwell L, et al. Interpretation of the evidence for the efficacy and safety of statin therapy. The Lancet. 2016;388:2532–61.

2. Cholesterol Treatment Trialists’ (CTT) Collaboration, Fulcher J, O’Connell R, Voysey M, Emberson J, Blackwell L, et al. Efficacy and safety of LDL-lowering therapy among men and women: meta-analysis of individual data from 174,000 participants in 27 randomised trials. Lancet. 2015;385:1397–405.

3. Visseren FLJ, Mach F, Smulders YM, Carballo D, Koskinas KC, Bäck M, et al. 2021 ESC Guidelines on cardiovascular disease prevention in clinical practice: Developed by the Task Force for cardiovascular disease prevention in clinical practice with representatives of the European Society of Cardiology and 12 medical societies With the special contribution of the European Association of Preventive Cardiology (EAPC). European Heart Journal. 2021. https://doi.org/10.1093/eurheartj/ehab484.

4. Mach F, Baigent C, Catapano AL, Koskinas KC, Casula M, Badimon L, et al. 2019 ESC/EAS Guidelines for the management of dyslipidaemias: lipid modification to reduce cardiovascular risk: The Task Force for the management of dyslipidaemias of the European Society of Cardiology (ESC) and European Atherosclerosis Society (EAS). European Heart Journal. 2020;41:111–88.

5. Arnett DK, Blumenthal RS, Albert MA, Buroker AB, Goldberger ZD, Hahn EJ, et al. 2019 ACC/AHA Guideline on the Primary Prevention of Cardiovascular Disease: A Report of the American College of Cardiology/American Heart Association Task Force on Clinical Practice Guidelines. Circulation. 2019;140.

6. Sommerschild H, Berg C, Blix H, Dansie L, Litleskare I, Olsen K, et al. Legemiddelforbruket i Norge 2016–2020. Data fra Grossistbasert legemiddelstatistikk og Reseptregisteret/ Drug Consumption in Norway 2016-2020 - Data from Norwegian Drug Wholesales Statistics and the Norwegian, Legemiddelstatistikk 2021. Oslo: Folkehelseinstituttet; 2021.

7. Reseptregisterets statistikkbank / Summary statistics from The Norwegian Prescription Database (NorPD). 2021.

8. Ference BA, Ginsberg HN, Graham I, Ray KK, Packard CJ, Bruckert E, et al. Low-density lipoproteins cause atherosclerotic cardiovascular disease. 1. Evidence from genetic, epidemiologic, and clinical studies. A consensus statement from the European Atherosclerosis Society Consensus Panel. Eur Heart J. 2017;38:2459–72.

9. Medisinsk fødselsregister statistikkbank / Summary statistics from the Medical Birth Registry of Norway (MBRN). 2021.

10. Irgens LM. [Medical birth registry--an essential resource in perinatal medical research]. Tidsskr Nor Laegeforen. 2002;122:2546–9.

11. Furu K, Wettermark B, Andersen M, Martikainen JE, Almarsdottir AB, Sørensen HT. The Nordic countries as a cohort for pharmacoepidemiological research. Basic Clin Pharmacol Toxicol. 2010;106:86–94.

12. Bateman BT, Mhyre JM, Hernandez-Diaz S, Huybrechts KF, Fischer MA, Creanga AA, et al. Development of a comorbidity index for use in obstetric patients. Obstet Gynecol. 2013;122:957–65.

13. R Core Team. R: A Language and Environment for Statistical Computing. Vienna, Austria; 2019.

14. Wickham H, Averick M, Bryan J, Chang W, McGowan L, François R, et al. Welcome to the Tidyverse. Journal of Open Source Software. 2019;4:1686.

15. Chang J-C, Chen Y-J, Chen I-C, Lin W-S, Chen Y-M, Lin C-H. Perinatal Outcomes After Statin Exposure During Pregnancy. JAMA Network Open. 2021;4:e2141321.

16. Lee M-S, Hekimian A, Doctorian T, Duan L. Statin exposure during first trimester of pregnancy is associated with fetal ventricular septal defect. Int J Cardiol. 2018;269:111–3.

17. Bateman BT, Hernandez-Diaz S, Fischer MA, Seely EW, Ecker JL, Franklin JM, et al. Statins and congenital malformations: cohort study. BMJ. 2015;350:h1035.

18. Colvin L, Slack-Smith L, Stanley FJ, Bower C. Linking a pharmaceutical claims database with a birth defects registry to investigate birth defect rates of suspected teratogens. Pharmacoepidemiol Drug Saf. 2010;19:1137–50.

19. Zomerdijk IM, Ruiter R, Houweling LMA, Herings RMC, Straus SMJM, Stricker BH. Dispensing of potentially teratogenic drugs before conception and during pregnancy: a population-based study. BJOG. 2015;122:1119–29.

20. Rachamin Y, Grischott T, Rosemann T, Meyer MR. Inferior control of low-density lipoprotein cholesterol in women is the primary sex difference in modifiable cardiovascular risk: A large-scale, cross-sectional study in primary care. Atherosclerosis. 2021;324:141–7.

21. Farukhi ZM, Mora S. Misperceptions and management of risk: Ongoing challenges in women’s cardiovascular health. Atherosclerosis. 2021;324:109–11.

22. Aceti A, Santhakumaran S, Logan KM, Philipps LH, Prior E, Gale C, et al. The diabetic pregnancy and offspring blood pressure in childhood: a systematic review and meta-analysis. Diabetologia. 2012;55:3114–27.

23. Cheung NW, Byth K. Population health significance of gestational diabetes. Diabetes Care. 2003;26:2005–9.

24. Cannon CP, Blazing MA, Giugliano RP, McCagg A, White JA, Theroux P, et al. Ezetimibe Added to Statin Therapy after Acute Coronary Syndromes. New England Journal of Medicine. 2015;372:2387–97.

25. Sakshaug S, Furu K, Karlstad Ø, Rønning M, Skurtveit S. Switching statins in Norway after new reimbursement policy: a nationwide prescription study. Br J Clin Pharmacol. 2007;64:476–81.

26. Steinberg D. Earlier intervention in the management of hypercholesterolemia: what are we waiting for? J Am Coll Cardiol. 2010;56:627–9.

27. Ference BA, Graham I, Tokgozoglu L, Catapano AL. Impact of Lipids on Cardiovascular Health: JACC Health Promotion Series. Journal of the American College of Cardiology. 2018;72:1141–56.

28. Langslet G, Holven KB, Bogsrud MP. Treatment goals in familial hypercholesterolaemia— time to consider low-density lipoprotein-cholesterol burden. European Journal of Preventive Cardiology. 2021;:zwab228.

29. Helsedirektoratet. Nasjonal faglig retningslinje for svangerskapsomsorgen / Norwegian national guideline for maternity care. Oslo: Helsedirektoratet; 2018.

30. The HAPO Study Cooperative Research Group. Hyperglycemia and Adverse Pregnancy Outcomes. New England Journal of Medicine. 2008;358:1991–2002.

31. Hafferty JD, Campbell AI, Navrady LB, Adams MJ, MacIntyre D, Lawrie SM, et al. Self-reported medication use validated through record linkage to national prescribing data. J Clin Epidemiol. 2018;94:132–42.

32. Herings R, Pedersen L. Pharmacy-based medical record linkage systems. In: Strom B, Kimmel S, Hennessy S, editors. Pharmacoepidemiology. 2012. p. 270–86.

