## Supplementary material for "Use of statins and other lipid-modifying agents across pregnancy – a nationwide drug utilization study of 822 071 pregnancies in Norway in 2005-2018": Online supplementary material

### Supplementary Results and Discussion

#### *Main statin types*

From *one year before conception* to *one year postpartum*, of all dispensations for lipid-modifying agents, one third was simvastatin and one third was atorvastatin (data not shown). For simvastatin, 20 and 40 mg/d were most commonly prescribed, whereas for atorvastatin it was also common to prescribe both 80 mg/d and 10 mg/d. Rosuvastatin was often prescribed in lower doses, including 5 and 10 mg/d. Statins were most widely prescribed in packages of 98 or 100 tablets, thereby covering over three month periods, assuming one tablet per day. Rosuvastatin was also prescribed for shorter periods, such as one month. These results correspond well with use of statin type and dose in clinical trials [1, 2]. Medium-intensity statins, particularly simvastatin, was the common choice previously; however, in recent years, high-intensity statins, particularly atorvastatin, has become more common (Figure 3). Rosuvastatin can also be considered high-intensity, even at low doses, since its LDL-C-lowering effect at 10-20 mg/d is approximately similar to atorvastatin at 40-80 mg/d [3]. Differences in solubility might also explain trends in popularity: while both simvastatin and atorvastatin are highly lipophilic statins, rosuvastatin is less lipophilic.

The different types and doses for statin prescription fills were evenly spread out before, during and after pregnancy, and there was no sign that pregnancy induced a shift from one statin type and dose to another (Figure S2).

#### *Characteristics of LMA users*

##### Non-statin LMA use

Non-statin LMA use across pregnancy varied with several maternal characteristics, including higher age (40 vs. 31 % above 32 years), more frequent use of folate prior to pregnancy (38 vs. 28 %), and more co-morbidities such as chronic hypertension (2.2 vs. 0.5 %) and diabetes mellitus (7.7 vs. 3.5 %) (Table S2). Women using omega-3 had particularly differing characteristics: compared with any non-statin LMA use, they had higher age (51 vs. 40 % above 32 years), had higher pre-pregnancy weight (74 vs. 68 kg), had experienced a previous pregnancy loss (32 vs. 24 %), had more frequent pre-pregnant folate use (41 vs. 38 %), and considerably more co-morbidities (30 vs. 13 % had any chronic illness). These results emphasize that women who use non-statin LMAs generally have poorer health status compared with women who do not use non-statin LMAs.

##### Discontinuation of statin use

Compared to women who used statins in pregnancy, women who discontinued their statin use earlier (that is, more than three months before conception, or zero to three months before conception) had several characteristics that could reflect *pregnancy planning* (Table S3). They were more often younger (46 and 44 % above 32 years for those discontinuing more than three months before conception and zero to three months before conception, respectively, vs. 54 % above 32 years for those using statins in pregnancy), employed (84 and 83 % vs. 74 %), had experienced a previous pregnancy loss (29 and 22 % vs. 23 %), had more frequently used folate before pregnancy (37 and 33 % vs. 26 %), lower proportion of smokers (22 and 26 % vs. 26 %), and lower proportion of comorbidities, such as chronic hypertension (5.3 and 5.7 % vs. 10 %) and diabetes (18 and 15 % vs. 24 %). Interestingly, more women discontinued their statin use early for their second pregnancy (primiparous women) compared to other pregnancies (38 and 31 % vs. 30 %). This may possibly be explained by how aware women are they are pregnant, and that they are better informed in their second pregnancy.

Women who filled their first statin prescriptions *after* pregnancy appeared *more similar* to the continuers than the two discontinuer groups (Table S3). For example, they had a more similar age (51 % vs. 54 %), employment status (76 vs. 73 %), and proportion of pre-pregnant folate use (28 vs. 26 %). In drug safety analyses, one common approach to selection of comparator group is using those discontinuing their drug use [4, 5], but these data suggest that comparing continuers with those starting immediately following pregnancy may be viable alternative.

Note also that many women did not start up their statin use immediately after pregnancy. A prescription for a statin was filled within one year before conception but not within one year after delivery for 59 % of all pregnancies (52 % of all women) (data not shown). Pregnancy has therefore induced significant off-statin periods for many women, which has resulted in higher plasma cholesterol and most likely accelerated atherosclerosis.

### History of statin use

Before pregnancy, a larger than expected number of women were long-term, experienced statin users (10-148 prescription fills), compared with intermediate-term users (4-9 prescription fills) and short-term users and beginners (1-3 prescription fills) (Figure S5). The drop in prescription fills the last three to six months before pregnancy occurred in all groups; also, all groups were represented as statin users in first trimester. After pregnancy, a larger than expected number of women were beginners. These data indicate that experienced statin users quit before pregnancy but then start up again rapidly, and that a proportion of women discover that they have elevated risk of ASCVD in pregnancy, possibly because of appropriate risk assessment, and then initiate statin therapy.

History of statin use prior to pregnancy also varied with many characteristics (Table S4). At time of conception, compared with short-term users and beginners, long-term, experienced statin users had higher proportion of employment (88 % vs. 81 %), nulliparity (50 vs. 38 %), folate use before pregnancy (40 vs. 32 %), and lower pre-pregnancy body weight (67 vs. 72 kg), lower proportion of smoking (15 vs. 29 %) and less chronic illness (23 vs. 27 %). Importantly, these results suggest that history of statin use may represent another sensitivity analysis in downstream drug safety analyses.

### Dyslipidemia diagnoses and LMA use

Among 7 817 pregnancies (covering 5 531 unique women) where the woman was registered with a dyslipidemia diagnosis (ICPC-2 code T93 in KUHR) at any time before pregnancy until one year after delivery, in 1 801 pregnancies (22.7 %, covering 1 254 unique women), the woman used LMA within one year before conception and one year after delivery. Among pregnancies where a dyslipidemia diagnosis was identified, compared with non-LMA users, LMA users generally smoked more frequently (21 vs. 17 %), and had higher proportion of chronic hypertension (4.7 vs. 1.7 %) and diabetes (11 vs. 7.8 %) on top of their dyslipidemia (Table S5). These results suggest that a large proportion of young women with dyslipidemia remain untreated with statins, for unknown reasons. One possibility is that they prefer treatment with diet and lifestyle over medication use.

### Indication for treatment with LMAs based on cardiovascular co-medication and diagnoses as a measure of co-morbidity

About 86 % of all LMA users had at least one indication for LMA (Figure S6), based on cardiovascular co-medication (drugs used in diabetes [ATC code A10], antithrombotic agents [B01], and cardiovascular system [C other than C10]) and relevant diagnoses (Table S1). A total of 953 pregnancies had exactly one unique LMA indication, whereas some pregnancies had up to 20 unique LMA indications. The number of unique LMA indications was related to the characteristics of the LMA users, and in general, the more indications, the more co-morbidity (Table S6). This was likely related to the higher age (53 vs. 39 above 32 years for 4-8 vs. zero indications for LMA therapy) and weight (78 vs. 64 kg). The number of LMA indications was also related to the established comorbidity index (26 vs. 3.4 % had a score of three or more). These data indicate that number of LMA indications (here defined as relevant co-medication and diagnoses) adequately capture information about disease severity related to use of statins and other LMAs.

### Recurrent pregnancies and statin use

Pregnancy-related statin use in the first pregnancy preceded use of statins in a second, recurrent pregnancy (Figure S7). A total of 245 women (0.052 %) used statin in their first pregnancy. Compared to those who did not use statins in their first pregnancy, fewer women went on to become pregnant again (32.7 % vs. 51.4 %), which may be related to pregnancy complications and perceived risk. Among those who did become pregnant again, almost 1 in 5 used statins also in their second pregnancy. Despite these figures, the absolute number of women using statins in both first and second pregnancy was low, which is reassuring. Recurrent use may generally be of minor importance; still, it seems clear that women who used statins in pregnancy should be provided information regarding pharmacotherapy and pregnancy planning.

**Supplementary References**

1. Mach F, Baigent C, Catapano AL, Koskinas KC, Casula M, Badimon L, et al. 2019 ESC/EAS Guidelines for the management of dyslipidaemias: lipid modification to reduce cardiovascular risk: The Task Force for the management of dyslipidaemias of the European Society of Cardiology (ESC) and European Atherosclerosis Society (EAS). European Heart Journal. 2020;41:111–88.

2. McKenney JM, Ganz P, Wiggins BS, Saseen JS. CHAPTER 22 - Statins. In: Ballantyne CM, editor. Clinical Lipidology. Philadelphia: W.B. Saunders; 2009. p. 253–80.

3. Jones PH, Davidson MH, Stein EA, Bays HE, McKenney JM, Miller E, et al. Comparison of the efficacy and safety of rosuvastatin versus atorvastatin, simvastatin, and pravastatin across doses (STELLAR* Trial). Am J Cardiol. 2003;92:152–60.

4. Hjorth S, Wood M, Tauqeer F, Nordeng H. Fertility treatment and oral contraceptive discontinuation for identification of pregnancy planning in routinely collected health data – an application to analgesic and antibiotic utilisation. BMC Pregnancy Childbirth. 2020;20:731.

5. Frank AS, Lupattelli A, Nordeng H. Risk factors for discontinuation of thyroid hormone replacement therapy in early pregnancy: a study from the Norwegian Mother and Child Cohort Study and the Medical Birth Registry of Norway. Acta Obstet Gynecol Scand. 2018;97:852–60.

### Supplementary Tables

#### *Table S1*

**Table S1.** Overview of diagnosis codes used as indications for lipid-modifying agent (LMA) use, as per Table S6 and Figure S6.

| System | Main group | Code | Disease |
|---|---|---|---|
| ICD-10 | E Endocrine, nutritional, metabolic | E10 | DM type I |
| ICD-10 | E Endocrine, nutritional, metabolic | E11 | DM type II |
| ICD-10 | E Endocrine, nutritional, metabolic | E12 | DM, undernutrition-based |
| ICD-10 | E Endocrine, nutritional, metabolic | E13 | DM, other |
| ICD-10 | E Endocrine, nutritional, metabolic | E14 | DM, unspecified |
| ICD-10 | I Circulatory | I10 | Essential (primary) hypertension |
| ICD-10 | I Circulatory | I11 | Hypertensive heart disease |
| ICD-10 | I Circulatory | I12 | Hypertensive kidney disease |
| ICD-10 | I Circulatory | I13 | Hypertensive heart-kidney disease |
| ICD-10 | I Circulatory | I15 | Secondary hypertension |
| ICD-10 | I Circulatory | I20 | Angina pectoris |
| ICD-10 | I Circulatory | I21 | Acute myocardial infarction |
| ICD-10 | I Circulatory | I25 | Chronic IHD |
| ICD-10 | I Circulatory | I26 | Lung embolus |
| ICD-10 | I Circulatory | I47 | Paroxysmal tachycardia |
| ICD-10 | I Circulatory | I49 | Heart arythmia |
| ICD-10 | I Circulatory | I50 | Heart failure |
| ICD-10 | I Circulatory | I60 | Subarachnoidal bleeding |
| ICD-10 | I Circulatory | I61 | Brain bleeding |
| ICD-10 | I Circulatory | I63 | Stroke |
| ICD-10 | I Circulatory | I65 | Stroke or bleeding, other |
| ICD-10 | I Circulatory | I67 | Brain disease, other |
| ICD-10 | I Circulatory | I69 | Disease caused by brain disease |
| ICD-10 | I Circulatory | I70 | Atherosclerosis |
| ICD-10 | I Circulatory | I72 | Aneurism/dissection, other |
| ICD-10 | I Circulatory | I73 | Peripheral vessel disease, other |
| ICD-10 | I Circulatory | I74 | Arterial embolus/thrombus |
| ICD-10 | I Circulatory | I82 | Venous embolus/thrombus |
| ICPC-2 | K Cardiovascular | K74 | IHD with angina pectoris |
| ICPC-2 | K Cardiovascular | K75 | Acute MI |
| ICPC-2 | K Cardiovascular | K76 | Chronic IHD |
| ICPC-2 | K Cardiovascular | K77 | Heart failure |
| ICPC-2 | K Cardiovascular | K80 | Arrythmia |
| ICPC-2 | K Cardiovascular | K81 | Murmur |
| ICPC-2 | K Cardiovascular | K84 | Heart disease |
| ICPC-2 | K Cardiovascular | K85 | Hypertension |
| ICPC-2 | K Cardiovascular | K86 | Hypertension |

| | | | |
|---|---|---|---|
| ICPC-2 | K Cardiovascular | K89 | TIA |
| ICPC-2 | K Cardiovascular | K90 | Stroke |
| ICPC-2 | K Cardiovascular | K91 | Cerebrovascular disease |
| ICPC-2 | K Cardiovascular | K92 | Atherosclerosis |
| ICPC-2 | K Cardiovascular | K93 | Lung embolus |
| ICPC-2 | K Cardiovascular | K99 | CVD |
| ICPC-2 | T Endocrine, metabolic, nutritional | T82 | Obesity |
| ICPC-2 | T Endocrine, metabolic, nutritional | T83 | Overweight |
| ICPC-2 | T Endocrine, metabolic, nutritional | T86 | Hypothyreosis |
| ICPC-2 | T Endocrine, metabolic, nutritional | T89 | DM type I |
| ICPC-2 | T Endocrine, metabolic, nutritional | T90 | DM type II |
| ICPC-2 | T Endocrine, metabolic, nutritional | T93 | Hyperlipidemia (incl. FH) |
| ICPC-2 | T Endocrine, metabolic, nutritional | T99 | Endocrine/metabolic/nutritional disease |

Abbreviations: CVD, cardiovascular disease; DM, diabetes mellitus; FH, familial hypercholesterolemia; ICD-10, the International Classification of Diseases, version 10; ICPC-2, the International Classification of Primary Care-2; IHD, ischemic heart disease; MI, myocardial infarction; TIA, transient ischemic attack.

*Table S2*

**Table S2.** Characteristics of the 822 071 pregnancies in 2005-2018, stratified by non-statin lipid-modifying agent (LMA) use between one year before conception and one year after delivery (covering 599 pregnancies and 519 unique women).

| | | No non-statin LMA use | Any non-statin LMA use | Specific non-statin LMA use | | |
|---|---|---|---|---|---|---|
| | n | n = 821 472 | n = 599 | Ezetimibe, n = 230 | Bile acid sequestrants, n = 311 | Omega-3, n = 73 |
| Age | 822 071 | | | | | |
| < 28 yrs | | 322 639 (39) | 195 (33) | 86 (37) | 98 (32) | 18 (25) |
| 28 - 32 yrs | | 240 746 (29) | 162 (27) | 62 (27) | 84 (27) | 18 (25) |
| > 32 yrs | | 258 087 (31) | 242 (40) | 82 (36) | 129 (41) | 37 (51) |
| Employed, yes | 656 916 | 531 345 (81) | 397 (81) | 177 (87) | 196 (78) | 37 (76) |
| Civil status | 822 071 | | | | | |
| Registered partner/cohabitant | | 762 311 (93) | 554 (92) | 217 (94) | 282 (91) | 69 (95) |
| Other | | 59 161 (7.2) | 45 (7.5) | 13 (5.7) | 29 (9.3) | 4 (5.5) |
| Parity | 822 071 | | | | | |
| Nulliparous | | 346 899 (42) | 274 (46) | 102 (44) | 145 (47) | 33 (45) |
| Primiparous | | 297 827 (36) | 207 (35) | 85 (37) | 103 (33) | 25 (34) |
| Multiparous | | 176 746 (22) | 118 (20) | 43 (19) | 63 (20) | 15 (21) |
| Previous pregnancy loss, yes | 771 869 | 182 785 (24) | 133 (24) | 44 (20) | 68 (24) | 22 (32) |
| Weight (pre-pregnancy), kg | 422 081 | 65 (58-75) | 68 (59-80) | 67 (58-76) | 70 (60-84) | 74 (60-86) |
| Folate use (pre-pregnancy), yes | 822 071 | 233 431 (28) | 227 (38) | 83 (36) | 117 (38) | 30 (41) |
| Smoking, yes | 552 527 | 107 568 (19) | 87 (20) | 25 (14) | 55 (24) | 7 (16) |
| Obstetric comorbidity index | 822 071 | | | | | |
| 0 | | 540 618 (66) | 328 (55) | 143 (62) | 171 (55) | 26 (36) |
| 1 | | 185 144 (23) | 171 (29) | 54 (23) | 94 (30) | 26 (36) |
| 2 | | 66 358 (8.1) | 51 (8.5) | 16 (7.0) | 26 (8.4) | 8 (11) |

| | | | | | | |
|---|---|---|---|---|---|---|
| 3 or more | | 29 352 (3.6) | 49 (8.2) | 17 (7.4) | 20 (6.4) | 13 (18) |
| Chronic illness, yes | 822 071 | 55 497 (6.8) | 80 (13) | 24 (10) | 35 (11) | 22 (30) |
| Chronic hypertension, yes | 822 071 | 4 431 (0.5) | 13 (2.2) | 5 (2.2) | 4 (1.3) | 4 (5.5) |
| Any diabetes mellitus, yes | 822 071 | 29 109 (3.5) | 46 (7.7) | 17 (7.4) | 15 (4.8) | 15 (21) |

n (%); Median (25%-75%).
Other non-statin LMAs were omitted because of low numbers (a total of n = 9 pregnancies).

*Table S3*

**Table S3.** Characteristics of 3 565 pregnancies (covering 2 608 unique women) in 2005-2018 where a statin prescription had been filled at any time before pregnancy until one year after delivery, stratified by discontinuation classification.

| | n | Discontinuation ≥ 3 months before conception, n = 2 045 | Discontinuation 0-3 months before conception, n = 566 | Statin use in pregnancy, n = 331 | Statin use initiated after pregnancy, n = 623 |
|---|---|---|---|---|---|
| Age | 3 565 | | | | |
| < 28 yrs | | 517 (25) | 155 (27) | 72 (22) | 163 (26) |
| 28 - 32 yrs | | 581 (28) | 161 (28) | 80 (24) | 140 (22) |
| > 32 yrs | | 947 (46) | 250 (44) | 179 (54) | 320 (51) |
| Employed, yes | 2 861 | 1 397 (84) | 389 (83) | 196 (73) | 353 (76) |
| Civil status | 3 565 | | | | |
| Registered partner/cohabitant | | 1 872 (92) | 522 (92) | 294 (89) | 559 (90) |
| Other | | 173 (8.5) | 44 (7.8) | 37 (11) | 64 (10) |
| Parity | 3 565 | | | | |
| Nulliparous | | 824 (40) | 251 (44) | 144 (44) | 206 (33) |
| Primiparous | | 783 (38) | 178 (31) | 98 (30) | 230 (37) |
| Multiparous | | 438 (21) | 137 (24) | 89 (27) | 187 (30) |
| Previous pregnancy loss, yes | 3 369 | 564 (29) | 120 (22) | 73 (23) | 172 (29) |
| Weight (pre-pregnancy), kg | 2 189 | 70 (61-82) | 70 (62-85) | 72 (61-85) | 73 (62-85) |
| Folate use (pre-pregnancy), yes | 3 565 | 755 (37) | 188 (33) | 87 (26) | 176 (28) |
| Smoking, yes | 2 675 | 346 (22) | 110 (26) | 65 (26) | 115 (29) |
| Obstetric comorbidity index | 3 565 | | | | |
| 0 | | 895 (44) | 265 (47) | 128 (39) | 224 (36) |
| 1 | | 582 (28) | 154 (27) | 86 (26) | 166 (27) |
| 2 | | 313 (15) | 76 (13) | 60 (18) | 130 (21) |
| 3 or more | | 255 (12) | 71 (13) | 57 (17) | 103 (17) |

| | | | | | |
|---|---|---|---|---|---|
| Chronic illness, yes | 3 565 | 505 (25) | 117 (21) | 109 (33) | 156 (25) |
| Chronic hypertension, yes | 3 565 | 109 (5.3) | 32 (5.7) | 34 (10) | 42 (6.7) |
| Any diabetes mellitus, yes | 3 565 | 377 (18) | 86 (15) | 80 (24) | 112 (18) |

n (%); Median (25%-75%)

*Table S4*

**Table S4**. Characteristics of 2 890 pregnancies (covering 2 089 unique women) in 2005-2018 where a statin prescription had been filled at any time before pregnancy until conception, stratified by history of statin use at time of the last prescription fill before conception.

| | n | 1-3 prescription fills, n = 1 370 | 4-9 prescription fills, n = 781 | 10-148 prescription fills, n = 739 |
|---|---|---|---|---|
| Year of delivery | 2 890 | | | |
| 2005 - 2009 | | 307 (22) | 171 (22) | 47 (6.4) |
| 2010 - 2014 | | 580 (42) | 335 (43) | 325 (44) |
| 2015 - 2018 | | 483 (35) | 275 (35) | 367 (50) |
| Age | 2 890 | | | |
| < 28 yrs | | 353 (26) | 207 (27) | 171 (23) |
| 28 - 32 yrs | | 359 (26) | 223 (29) | 226 (31) |
| > 32 yrs | | 658 (48) | 351 (45) | 342 (46) |
| Employed, yes | 2 356 | 888 (81) | 524 (82) | 546 (88) |
| Civil status | 2 890 | | | |
| Registered partner/cohabitant | | 1 231 (90) | 729 (93) | 679 (92) |
| Other | | 139 (10) | 52 (6.7) | 60 (8.1) |
| Parity | 2 890 | | | |
| Nulliparous | | 515 (38) | 322 (41) | 366 (50) |
| Primiparous | | 504 (37) | 295 (38) | 245 (33) |
| Multiparous | | 351 (26) | 164 (21) | 128 (17) |
| Previous pregnancy loss, yes | 2 739 | 370 (29) | 204 (27) | 165 (23) |
| Weight (pre-pregnancy), kg | 1 874 | 72 (62-84) | 70 (62-82) | 67 (60-80) |
| Folate use (pre-pregnancy), yes | 2 890 | 435 (32) | 283 (36) | 296 (40) |
| Smoking, yes | 2 237 | 296 (29) | 122 (21) | 94 (15) |

| | | | | |
|---|---|---|---|---|
| Obstetric comorbidity index | 2 890 | | | |
| 0 | | 558 (41) | 358 (46) | 351 (47) |
| 1 | | 388 (28) | 206 (26) | 212 (29) |
| 2 | | 230 (17) | 121 (15) | 88 (12) |
| 3 or more | | 194 (14) | 96 (12) | 88 (12) |
| Chronic illness, yes | 2 890 | 369 (27) | 177 (23) | 169 (23) |
| Chronic hypertension, yes | 2 890 | 82 (6.0) | 45 (5.8) | 43 (5.8) |
| Any diabetes mellitus, yes | 2 890 | 274 (20) | 138 (18) | 118 (16) |

n (%); Median (25%-75%)

*Table S5*

**Table S5.** Characteristics of 7 817 pregnancies (covering 5 531 unique women) in 2005-2018 where a diagnosis of dyslipidemia (ICPC-2 code T93) was recorded at any time before pregnancy until one year after delivery, stratified by any lipid-modifying agent (LMA) use (both statin and non-statin types) between one year before pregnancy and one year after delivery.

| | n | Dyslipidemia, but no LMA use across pregnancy, n = 6 016 | Dyslipidemia and LMA use across pregnancy, n = 1 801 |
|---|---|---|---|
| Age | 7 817 | | |
| < 28 yrs | | 1 681 (28) | 541 (30) |
| 28 - 32 yrs | | 1 824 (30) | 519 (29) |
| > 32 yrs | | 2 511 (42) | 741 (41) |
| Employed, yes | 6 420 | 4 247 (86) | 1 248 (84) |
| Civil status | 7 817 | | |
| Registered partner/cohabitant | | 5 641 (94) | 1 663 (92) |
| Other | | 375 (6.2) | 138 (7.7) |
| Parity | 7 817 | | |
| Nulliparous | | 2 429 (40) | 724 (40) |
| Primiparous | | 2 296 (38) | 666 (37) |
| Multiparous | | 1 291 (21) | 411 (23) |
| Previous pregnancy loss, yes | 7 455 | 1 502 (26) | 441 (26) |
| Weight (pre-pregnancy), kg | 5 070 | 69 (61-81) | 69 (61-82) |
| Folate use (pre-pregnancy), yes | 7 817 | 2 228 (37) | 617 (34) |
| Smoking, yes | 5 969 | 765 (17) | 293 (21) |
| Obstetric comorbidity index | 7 817 | | |
| 0 | | 3 237 (54) | 944 (52) |
| 1 | | 1 651 (27) | 480 (27) |

| | | | |
|---|---|---|---|
| 2 | | 728 (12) | 217 (12) |
| 3 or more | | 400 (6.6) | 160 (8.9) |
| Chronic illness, yes | 7 817 | 609 (10) | 278 (15) |
| Chronic hypertension, yes | 7 817 | 100 (1.7) | 85 (4.7) |
| Any diabetes mellitus, yes | 7 817 | 467 (7.8) | 207 (11) |

n (%); Median (25%-75%)

*Table S6*

**Table S6**. Characteristics of 2 772 pregnancies (covering 2 304 unique women) in 2005-2018 where a lipid-modifying agent (LMA) prescription had been filled between one year before pregnancy until one year after delivery, stratified by the number of unique drug and diagnosis indications for LMA therapy.

| | | Number of indications for LMA therapy | | | | |
|---|---|---|---|---|---|---|
| | | 0 | 1 | 2 - 3 | 4 - 8 | ≥ 9 |
| | n | n = 384 | n = 953 | n = 664 | n = 619 | n = 152 |
| Age | 2 772 | | | | | |
| < 28 yrs | | 112 (29) | 314 (33) | 179 (27) | 143 (23) | 15 (9.9) |
| 28 - 32 yrs | | 124 (32) | 266 (28) | 166 (25) | 145 (23) | 32 (21) |
| > 32 yrs | | 148 (39) | 373 (39) | 319 (48) | 331 (53) | 105 (69) |
| Employed, yes | 2 211 | 241 (82) | 662 (85) | 445 (82) | 355 (73) | 79 (71) |
| Civil status | 2 772 | | | | | |
| Registered partner/cohabitant | | 354 (92) | 883 (93) | 607 (91) | 550 (89) | 129 (85) |
| Other | | 30 (7.8) | 70 (7.3) | 57 (8.6) | 69 (11) | 23 (15) |
| Parity | 2 772 | | | | | |
| Nulliparous | | 181 (47) | 404 (42) | 265 (40) | 246 (40) | 56 (37) |
| Primiparous | | 126 (33) | 345 (36) | 214 (32) | 202 (33) | 53 (35) |
| Multiparous | | 77 (20) | 204 (21) | 185 (28) | 171 (28) | 43 (28) |
| Previous pregnancy loss, yes | 2 612 | 79 (23) | 201 (22) | 158 (26) | 191 (32) | 51 (36) |
| Weight (pre-pregnancy), kg | 1 558 | 64 (58-75) | 66 (58-76) | 71 (62-85) | 78 (63-93) | 84 (72-97) |
| Folate use (pre-pregnancy), yes | 2 772 | 150 (39) | 317 (33) | 232 (35) | 192 (31) | 46 (30) |
| Smoking, yes | 2 027 | 60 (28) | 152 (21) | 128 (25) | 128 (28) | 32 (26) |
| Obstetric comorbidity index | 2 772 | | | | | |
| 0 | | 239 (62) | 597 (63) | 288 (43) | 103 (17) | 8 (5.3) |
| 1 | | 109 (28) | 231 (24) | 207 (31) | 193 (31) | 23 (15) |
| 2 | | 23 (6.0) | 87 (9.1) | 93 (14) | 159 (26) | 45 (30) |
| 3 or more | | 13 (3.4) | 38 (4.0) | 76 (11) | 164 (26) | 76 (50) |

| | | | | | | |
|---|---|---|---|---|---|---|
| Chronic illness, yes | 2 772 | 26 (6.8) | 61 (6.4) | 103 (16) | 338 (55) | 109 (72) |
| Chronic hypertension, yes | 2 772 | 0 (0) | 4 (0.4) | 16 (2.4) | 84 (14) | 60 (39) |
| Any diabetes mellitus, yes | 2 772 | 3 (0.8) | 28 (2.9) | 78 (12) | 256 (41) | 89 (59) |

n (%); Median (25%-75%)

### Supplementary Figures

*Figure S1*

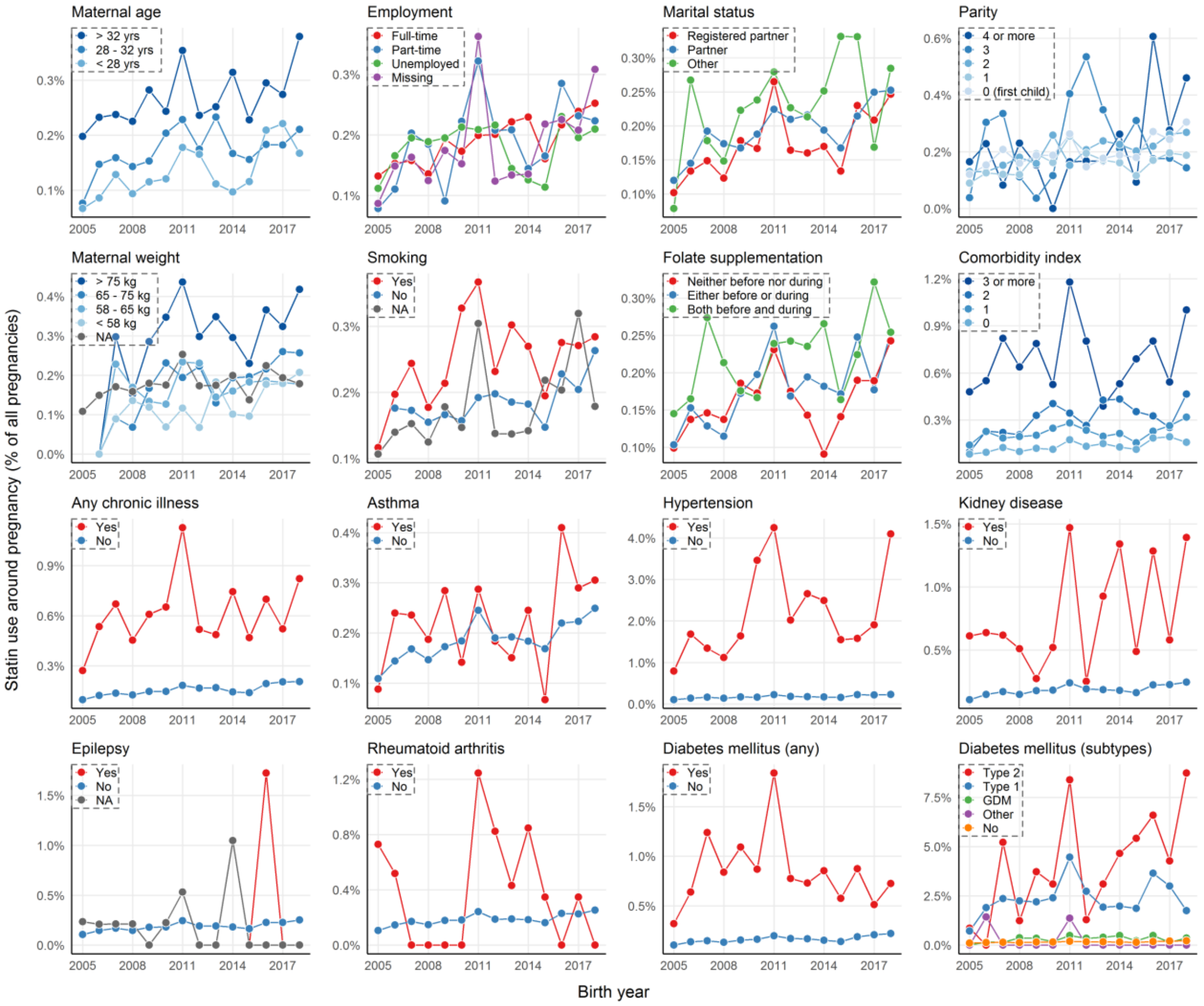


**Figure S1**. *Temporal trends in use of LMAs around pregnancy, stratified by maternal characteristics*. The figure contains data on those that filled a prescription for a lipid-modifying agent (LMA) within three months prior to pregnancy or during the entirety of pregnancy (any trimester) in Norway in 2005 to 2018. Maternal characteristics were taken from The Medical Birth Registry of Norway (MBRN).

*Figure S2*

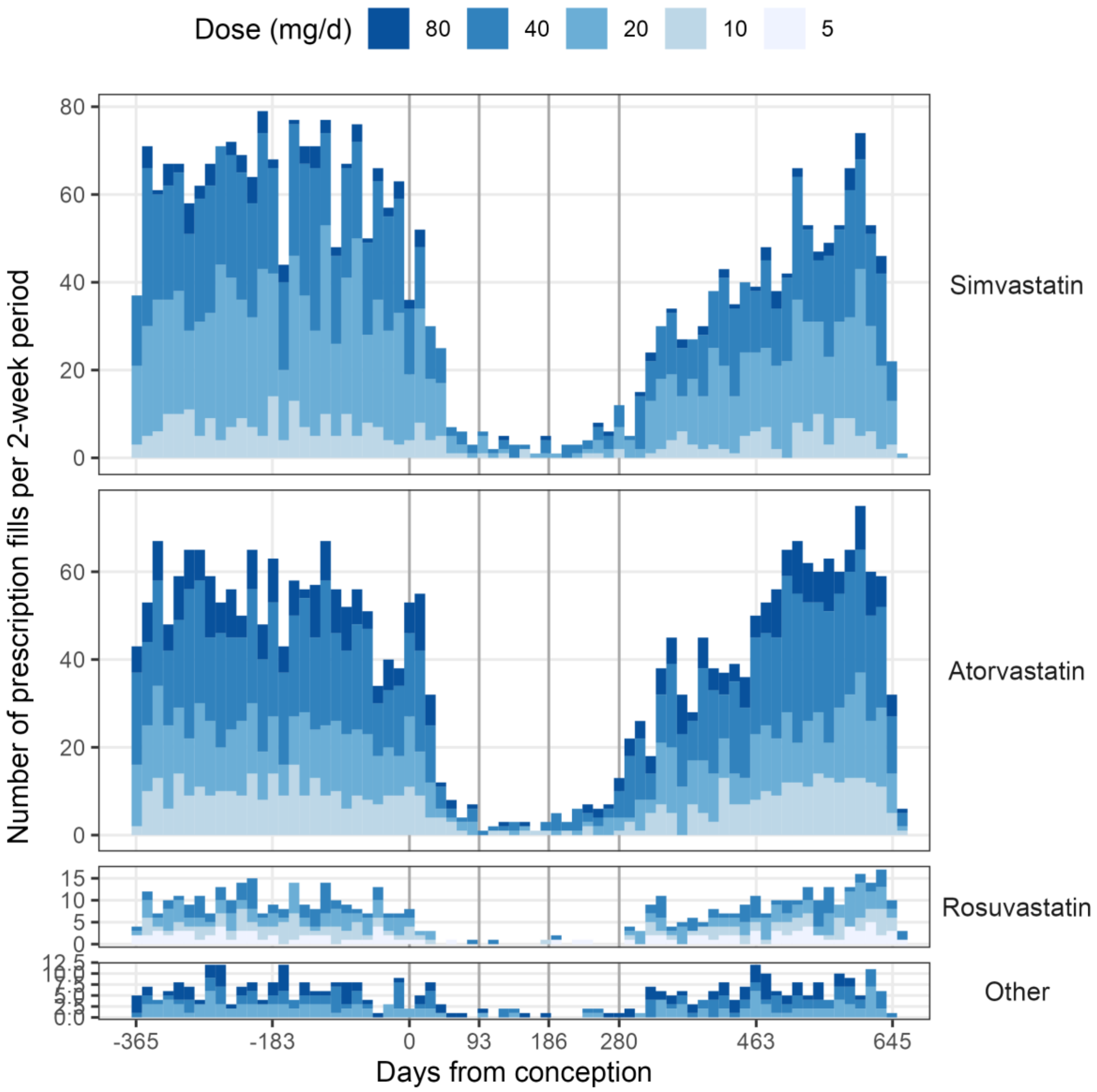


**Figure S2**. *Pattern of statin use one year before, during, and one year after pregnancy: variation in dose*. The figure shows 6 703 prescription fills for all women in Norway that were pregnant in the period 2005 to 2018 and that filled a prescription for a statin (ATC groups C10AA and C10B) between one year before conception and one year after delivery (2 439 pregnancies, 2 010 unique women). Data are shown in 2-week bins, stratified by statin type, and colored by statin dose. The vertical lines represent key timepoints in relation to time of conception, including conception and trimester transitions.

*Figure S3*

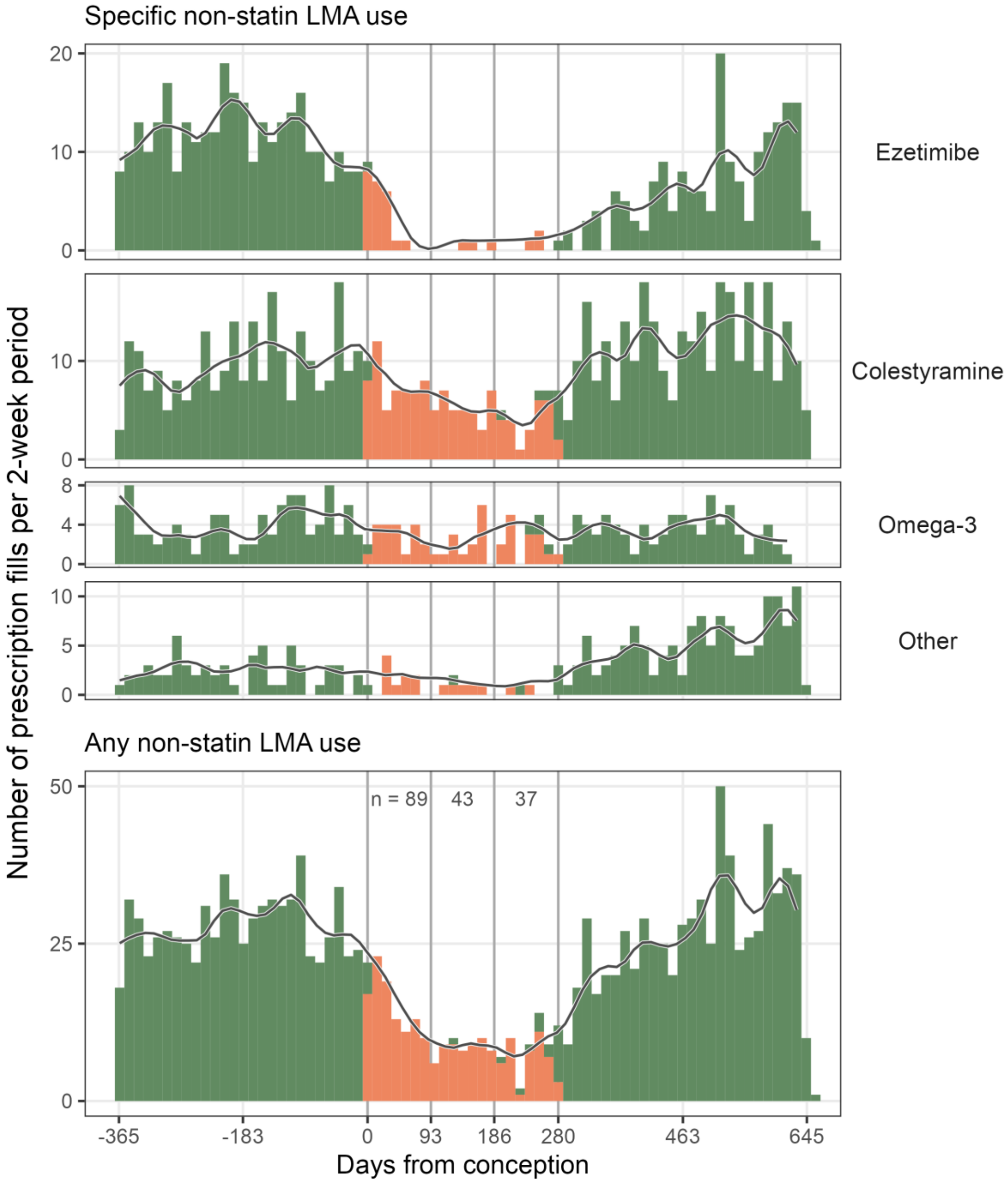


**Figure S3**. *Pattern of prescription fills for non-statin LMAs, one year before, during, and one year after pregnancy*. The figure shows 1 689 prescription fills for all women in Norway that were pregnant in the period 2005 to 2018 and that filled a prescription for a non-statin LMA (ATC group C10 except C10AA and C10B) between one year before conception and one year after delivery (610 pregnancies, 526 unique women). The figure shows the number of prescriptions fills of non-statin LMAs in 2-week bins, as well as a smoothened 2-month average (one month prior and the subsequent two weeks; local regressions were fit on 10 % of the data points along the line). Green and red color represents prescription fill of a statin *outside* or *within* any trimester of pregnancy, respectively. Numbers between the vertical lines in the upper part of the lowest panel represent the number of unique pregnancies where a statin prescription had been filled inside pregnancy, per trimester. The vertical lines represent key timepoints in relation to time of conception, including conception and trimester transitions.

*Figure S4*

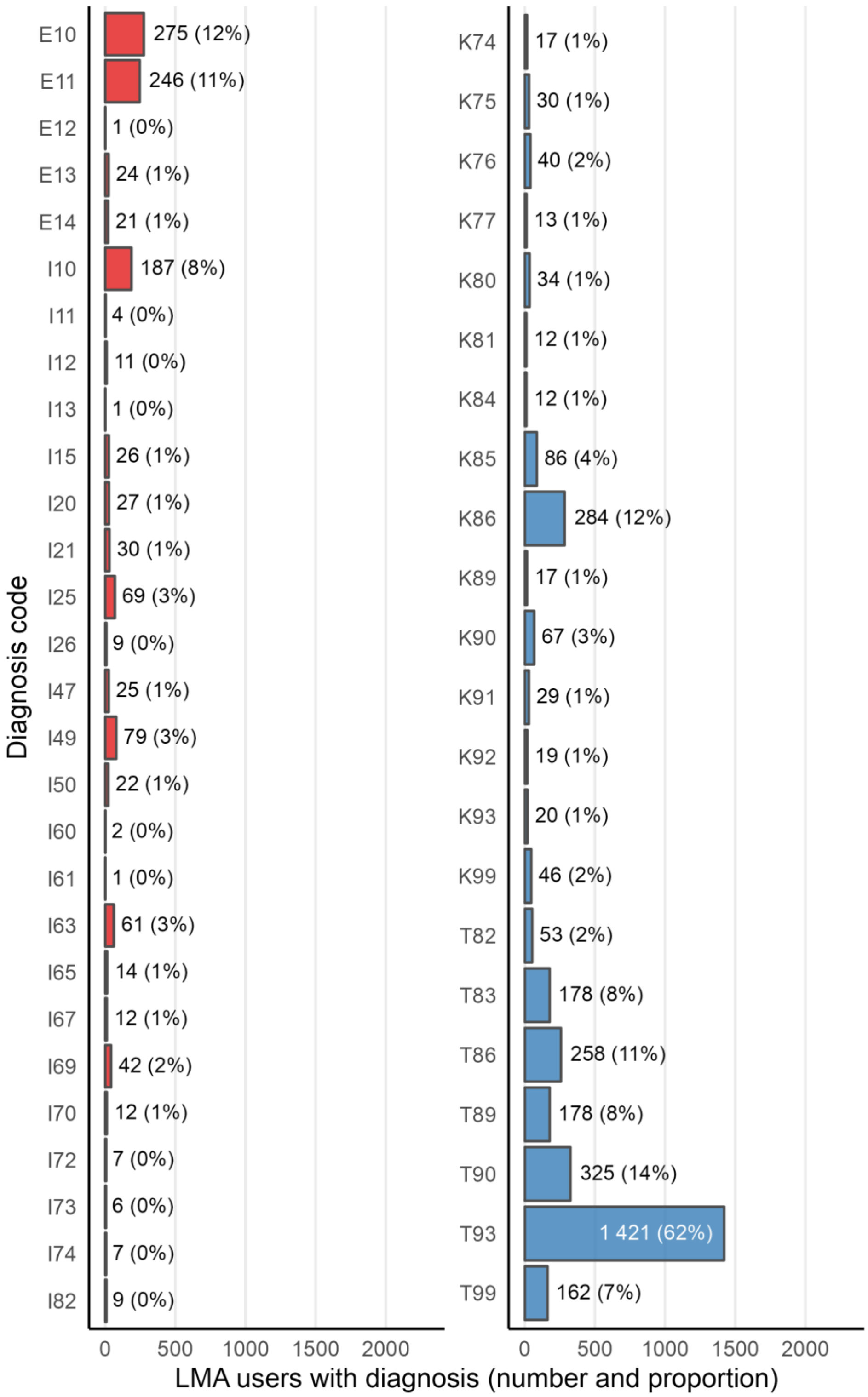


**Figure S4**. *Presence of relevant diagnoses among pregnancies where a prescription for an LMA had been filled*. The figure shows the number of pregnancies where a prescription for any LMA had been filled between one year before and one year after pregnancy, and where the woman had a diagnosis pertinent to LMA use at any timepoint before one year after delivery. Red is ICD-10 codes (either NPR or KUHR), and blue is ICPC-2 codes (KUHR only).

The diagnosis groups are in alphabetical order. The x axis has been scaled to the total number of unique pregnancies (n = 2 439), and the percentages reflect the proportion of all unique pregnancies where the particular diagnosis was identified. Diagnosis codes are annotated in Table S1; also, see https://finnkode.ehelse.no/ for more information about the codes (website in Norwegian). Abbreviations: ICD-10, The International Classification of Diseases, version 10; ICPC-2, The International Classification of Primary Care-2; KUHR, The Norway Control and Payment of Health Reimbursement Database; LMA, lipid-modifying agents; NPR, The Norwegian Patient Registry.

*Figure S5*

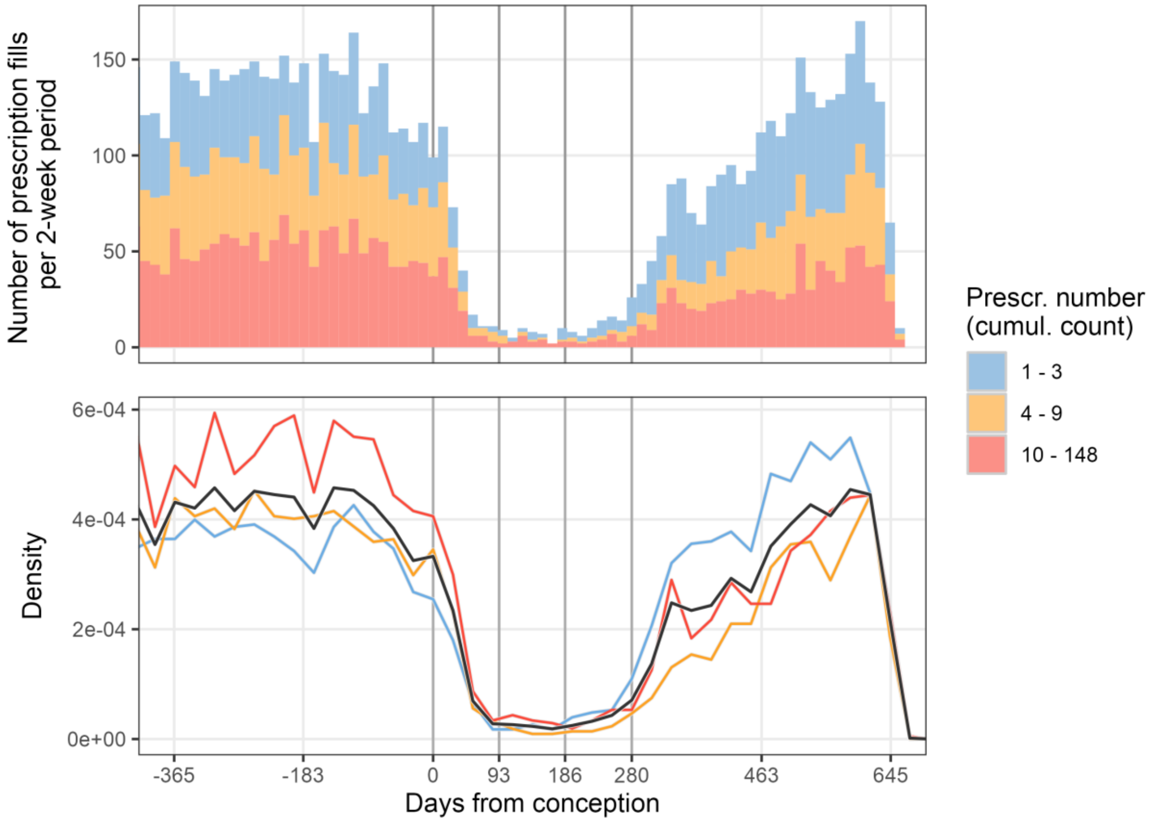


**Figure S5**. *Pattern of history of statin use one year before, during, and one year after pregnancy, stratified by history of statin use at a given point in time*. The figure shows history of statin use across pregnancy; history of statin use was calculated as the chronological, rolling count of prescription fills per woman and pregnancy. We determined the ranges by binning the number of prescriptions into three groups of approximately equal number of observations. The histogram shows 2-week bins, and the frequency polygons shows 1-month bins. While the colors denote groups of prescription fill numbers (per woman and pregnancy), the black line represents the average ("expected") numbers. The numbers for the frequency polygons are on *density* scale, the values of which are uninterpretable; interpret the colored groups *in relation to the black line*. The vertical lines represent key timepoints in relation to time of conception, including conception and trimester transitions. Note that this figure shows the *running* history of statin use, while Table S4 shows the last history of statin use before conception.

*Figure S6*

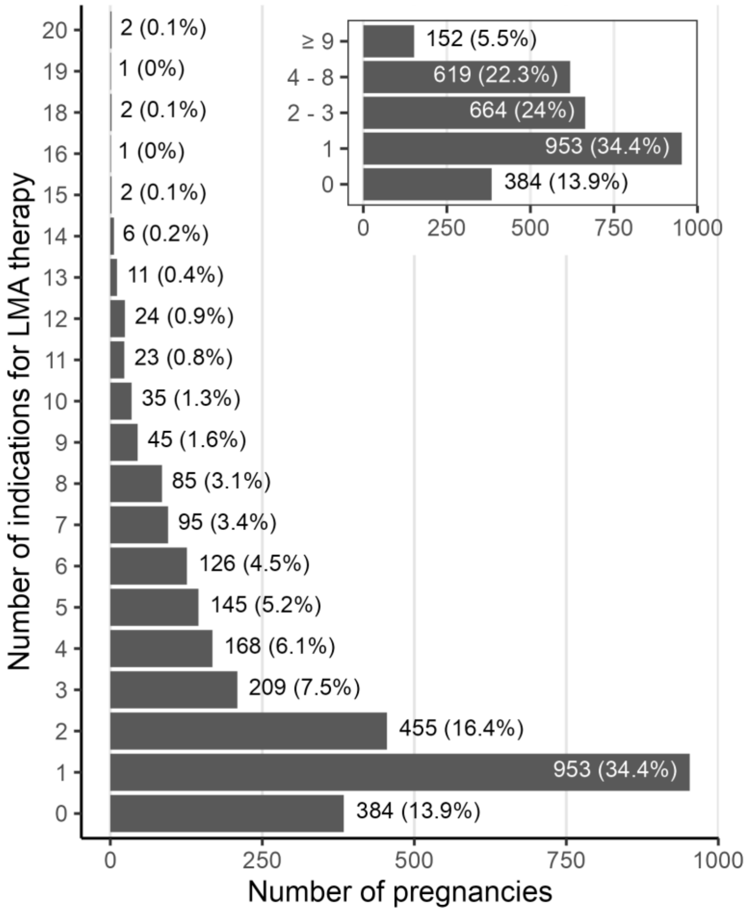


**Figure S6**. *Co-medication and diagnoses as indications for LMA use*. The figure shows the number of unique indications for LMA use per woman and pregnancy. Indications were defined as the sum of drugs used in diabetes (ATC code A10), antithrombotic agents (B01), and cardiovascular system (C other than C10), and all diagnosis codes defined in Table S1, per woman and pregnancy. The inset is a binned representative of the main panel, which show the actual values. Abbreviations: ATC, The World Health Organization's Anatomical Therapeutic Chemical Classification System; LMA, lipid-modifying agents.

*Figure S7*

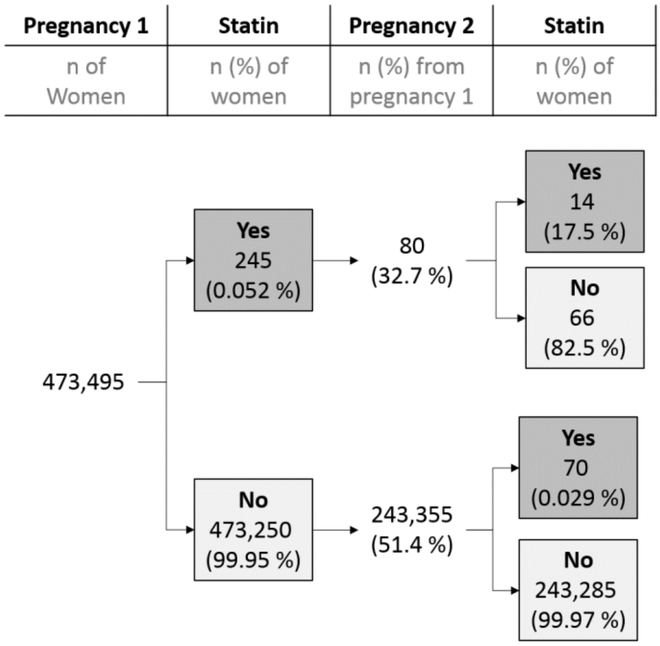


**Figure S7**. *Statin use in a recurrent pregnancy*. The figure shows statin prescription fills in a given pregnancy, split by pregnancy order and statin prescription fills. Only the first two pregnancies are shown due to low counts in subsequent pregnancies.
